# Estimating the changing prevalence of molecular markers of artemisinin partial resistance in *Plasmodium falciparum* malaria in Sub-Saharan Africa

**DOI:** 10.64898/2026.03.03.26347488

**Authors:** Lucinda E. Harrison, Nick Golding, Tianxiao Hao, Imke Botha, Stephanie van Wyk, Donnie Mategula, Prabin Dahal, Jaishree Raman, Daniel J. Weiss, Karen I. Barnes, Philippe J. Guérin, Jennifer A. Flegg

## Abstract

**Background:** Artemisinin-based combination therapies (ACTs) are the most widely used treatment for *Plasmodium falciparum* malaria. Kelch 13 mutations associated with artemisinin partial resistance (ART-R) have emerged in Sub-Saharan Africa (SSA) and are now reported in an increasing number of countries. ACT treatment failure rates are at risk of unprecedented increase. To summarise existing surveillance data and guide future surveillance, we produce modelled estimates of the spatiotemporal distributions of Kelch 13 and partner drug marker prevalence in SSA.

**Methods:** We develop and validate spatiotemporal statistical models, fitted within a Bayesian framework, given molecular surveillance data. We estimate the prevalence of Kelch 13 mutations that are validated or candidate markers of ART-R and the prevalence of the mutations *Pfcrt*-K76T, *Pfmdr1*-N84Y, *Pfmdr1*-Y186F, and *Pfmdr1*-D1246Y, associated with selection by pressure from the ACT partner drugs amodiaquine and lumefantrine.

**Findings:** Our models reflect all existing clusters of ART-R-associated Kelch 13 mutations. We estimate the prevalence of these Kelch 13 mutations to be greater than 10% in 23% of the area of endemic malaria transmission in SSA in 2026. We also estimate that 5.8% of malaria cases in 2026 will be affected by a validated or a candidate ART-R marker. Our estimates of the prevalence of *Pfcrt*-K76T and other partner drug markers reflect sustained pressure from artemether-lumefantrine: we estimate the median prevalence of *Pfcrt*-76T across SSA to be 19% in 2026.

**Interpretation:** Our models allow readers to visualise variation in observed mutation prevalences and to extrapolate prevalence to regions in space and time that are not represented in surveillance data. To monitor the changing distribution of antimalarial resistance markers within the constraints of the current global health funding climate it is critical that validated, statistical frameworks are incorporated into decision-making workflows to make the best use of molecular surveillance data.

**Funding:** This research was funded by the European Union under the Global Health EDCTP3 Joint Undertaking (grant agreement 101103076) and the Australian National Health and Medical Research Council (APP2019093).

**Research in context:** *Evidence before this study:* Ongoing systematic reviews of molecular surveillance of antimalarial resistance markers collate evidence of changing prevalences of Kelch 13 mutations associated with artemisinin partial resistance. However, there is a high degree of sampling bias in this data, and there are regions where limited surveillance has been carried out. We searched PubMed with the search terms: (((spatial OR spatiotemporal) AND (artemisinin OR Kelch)) AND (Africa)) AND (model* OR map OR mapping) which returned 30 results. We identified one recent pre-print describing spatiotemporal models of molecular markers of ART-R and partner drug resistance, however these models were not formally validated and model uncertainty may have been under-estimated.

*Added value of this study:* We use spatiotemporal statistical models to estimate resistance marker prevalence in regions where there has been no molecular surveillance. Our models’ predictions are contextualised by estimates of model uncertainty, and we validate our modelling framework through posterior predictive checks and by evaluating its predictive performance on held-out data.

*Implications of all available evidence:* Kelch 13 mutation prevalences are rising in all existing clusters where mutations have been identified, including in southern Africa. We estimate elevated prevalences in regions that neighbour existing clusters that are not well-represented in our surveillance dataset.

## 1 Introduction

With more than 280 million reported cases and 600,000 deaths in 2024, malaria remains a significant burden for the poorest countries of the world [1]. Approximately 94% of malaria cases globally occur in Africa, and of these, the vast majority are caused by the parasite *Plasmodium falciparum* [1]. Progress towards malaria elimination has stagnated in the last decade, with drug resistance being a key contributor to the situation [1].

Since the turn of the 21st century, global antimalarial pressure has shifted to artemisinin combination therapies (ACTs) [2, 3, 4]. An ACT is a combination of a fast-acting drug (the artemisinin derivatives) with a longer-lasting partner drug, such as lumefantrine (as artemether-lumefantrine; AL), amodiaquine (as artesunate-amodiaquine; ASAQ), piperaquine (as dihydroartemisinin-piperaquine; DHA-PPQ) or pyronari-dine (artesunate-pyronarydine; APyr) [3, 5]. ACTs were first introduced at scale in the 1990s in South East Asia after the spread of parasite resistance to chloroquine and its replacement, sulfadoxine-pyrimethamine (SP), and the subsequent rise in treatment failures [3, 4, 6]. The first instances of reduced efficacy of ACTs were reported in the Greater Mekong Subregion (GMS) in 2008 [7, 8], in the form of delayed parasite clearance attributed to resistance to the artemisinin component of the drug combination. Artemisinin partial resistance (ART-R) was later experimentally linked to parasite mutations in the propeller region of the *Pfkelch13* gene [9]. The World Health Organization (WHO) currently identifies 20 validated and candidate markers of ART-R in the *Pfkelch13* gene [10]. By 2022, the prevalence of ART-R-associated Kelch 13 mutations was close to fixation throughout *P. falciparum* populations in Cambodia, southern Laos, and malaria-endemic areas in southern Vietnam; the dominant mutant across the region is C580Y [11, 12]. ART-R-associated Kelch 13 mutations emerged independently in the Great Lakes region of East Africa and in the Horn of Africa as early as 2015 [13, 14, 15].

Molecular markers of resistance to some ACT partner drugs have also been identified. By 2001, the point mutations *Pfcrt*-76T and *Pfmdr1*-86Y were linked to chloroquine resistance [16]. Subsequently, the *Pfmdr1*-86Y-Y184-1246Y (YYY) haplotype, and its inverse *Pfmdr1*-N86-184F-D1246 (NFD), were associated with parasite recrudescence following treatment with ASAQ and AL, respectively [17, 18, 19]. *Pfmdr1*-N86 was independently associated with parasite recrudescence following treatment with AL [20]. While *Pfcrt*-76T was prevalent in Sub-Saharan Africa after decades of extensive chloroquine use, prevalence has dropped following widespread policy shifts to AL [21]. Increased prevalence of *Pfmdr1*-NFD has been found to coincide with national policies recommending first-line treatment with AL, while the same was true of *Pfmdr1*-YYY in countries which recommended ASAQ [22].

There are several recent reviews of Kelch 13 surveillance efforts in Africa [5, 23, 24]. Contributing towards each of these reviews, the Worldwide Antimalarial Resistance Network (WWARN) Molecular Surveyor dataset is a longstanding living systematic review of published or pre-print records of molecular surveillance in *P. falciparum* malaria [25]. The WWARN Molecular Surveyor collates records of Kelch 13 mutation surveillance, as well as surveillance of other established markers of resistance to ACTs and other antimalarials, including the *Pfdhps* and *Pfdhfr* markers associated with SP resistance and putative markers for reduced susceptibility to ASAQ and AL in the *Pfcrt* and *Pfmdr1* genes [26]. The Surveyor is routinely updated and is accessible via an interactive dashboard [25]. Drawing on the Surveyor, the MARC SE-Africa antimalarial resistance dashboard [5] further includes several recent pre-publication records of surveillance in Southern and Eastern Africa, including those in the WHO Malaria Threats Map dataset [27].

Monitoring the prevalence of molecular markers for drug resistance serves as an early warning system for increasing risks of treatment failure, and has the potential for far greater spatial coverage and recency than what could be feasibly included in therapeutic efficacy studies alone [28]. Molecular surveillance is particularly important where rising clinical ART-R is masked by partner drug efficacy [29]. Individual molecular surveillance efforts may be carefully designed in the context of specific hypothesis tests and prioritising specific markers and populations [30]. However, when aggregated to a national, continental, or global scale, existing *P. falciparum* molecular surveillance data are highly spatiotemporally biased and incomplete. This is generally true of disease surveillance data: it is not usually feasible to exhaustively survey an entire target population when surveillance resources are finite [31, 32]. It is therefore critical that molecular surveillance data is leveraged across national borders to provide a comprehensive picture of the current state of genetic diversity in the parasite population for all local decision-makers across malaria-endemic regions [30].

Geostatistical models enable the estimation of local molecular marker prevalences in places with no surveillance using nearby surveillance data, while accounting for sampling bias. They are widely employed to estimate the spatiotemporal distributions of molecular markers for antimalarial resistance as continuous maps, including Kelch 13 mutation prevalence in the GMS [11] and India [33], and markers for SP resistance in Africa [34, 35] and India [33]. Model-based mapping of Kelch 13 mutation prevalence in Africa is emerging in recent literature [24], however these models are not formally validated and the emerging cluster of mutants in Southern Africa (reported in both Namibia [36] and Zambia [37, 38]) has been neglected. Ultimately, the visualisation of spatiotemporal variation in uncertainty and model performance, alongside median point estimates, is critical to providing robust and interpretable results of geostatistical modelling to decision-makers [32].

In this work, we develop validated spatiotemporal models of the prevalence of Kelch 13 ART-R markers in Sub-Saharan Africa to inform policy-making. We also develop maps of four mutations in the *Pfcrt* and *Pfmdr1* genes, as markers of reduced parasite susceptibility to amodiaquine and lumefantrine, the most widely used ACT partner drugs in Sub-Saharan Africa. Alongside our models, we provide complete estimates of model uncertainty and model performance. By considering these models together, decision-makers are provided with a comprehensive estimate of the spatiotemporal landscape of parasite susceptibility to ACTs in Sub-Saharan Africa.

## 2 Methods

In this work we create ten spatiotemporal models of molecular markers for antimalarial resistance: firstly, an aggregate Kelch 13 model of the prevalence of *all* ART-R validated and candidate mutations (Table S1). While relationships between each mutation and the clinical response may be heterogeneous, this aggregate model shows high-level potential for ART-R across the region of endemic malaria transmission in Africa. Independently of the Kelch 13 aggregate model, we also model the five most frequently identified ART-R-associated Kelch 13 mutations in Africa: C469Y, A675V, R561H, P441L, and R622I. Finally, we model mutation prevalence at four loci putatively associated with reduced parasite susceptibility to the ACT partner drugs lumefantrine and amodiaquine: *Pfcrt*-K76T, *Pfmdr1*-N86Y, *Pfmdr1*-Y184F, and *Pfmdr1*-D1246Y. Wild-type and mutant alleles at the latter three of these loci jointly constitute the *Pfmdr1*-YYY and *Pfmdr1*-NFD haplotypes.

The molecular surveillance dataset we draw on in this work is the product of (1) the WWARN Molecular Surveyor [25, 39], a living systematic review of published and preprint records of molecular surveillance of *P. falciparum* and (2) the MARC SE-Africa antimalarial resistance dashboard [5], which complements the Surveyor by including pre-publication and unpublished records of molecular surveillance in Southern and Eastern Africa, including those displayed on the WHO Malaria Threats Map [27].

We model mutation prevalences using Gaussian Process models [40] with spatiotemporal Gneiting class kernels [35, 41]. Both time and modelled estimates of *Plasmodium falciparum* parasite rate [42] are included as model covariates. We assume mutation prevalences follow a beta-binomial distribution. While models of prevalence data are often assumed to follow a binomial distribution (e.g., [34, 35, 24]), the beta-binomial distribution includes an additional free parameter, compared to the binomial, that allows us to account for overdispersion empirically observed in molecular surveillance data. We fit the models using Bayesian inference and then use the fitted models to generate nominal 5×5 km^2^ annual regional maps of estimated drug resistance marker prevalences from 2000 to 2028. We validate our predictions by visualising coverage probabilities and performing posterior predictive checking. We evaluate residual errors using 10-fold stratified cross validation. We include complete descriptions of the molecular surveillance dataset, modelling framework, inference, and validation in the Supplementary Methods.

## 3 Results

### 3.1 Kelch 13

Recorded Kelch 13 surveillance intensity is greatest in Eastern Africa, in the Great Lakes region (Figures 1 and S1): 12% of all samples were collected in Uganda. Of the Kelch 13 mutations recognised as validated and candidate markers of ART-R (Table S1), the most frequently identified were C469Y, A675V, R561H, P441L, and R622I: the first four of these were often identified in the Great Lakes region. Elsewhere, R622I was primarily reported in the Horn of Africa and P441L predominated in Southern Africa (Figure 1).

**Figure 1.**
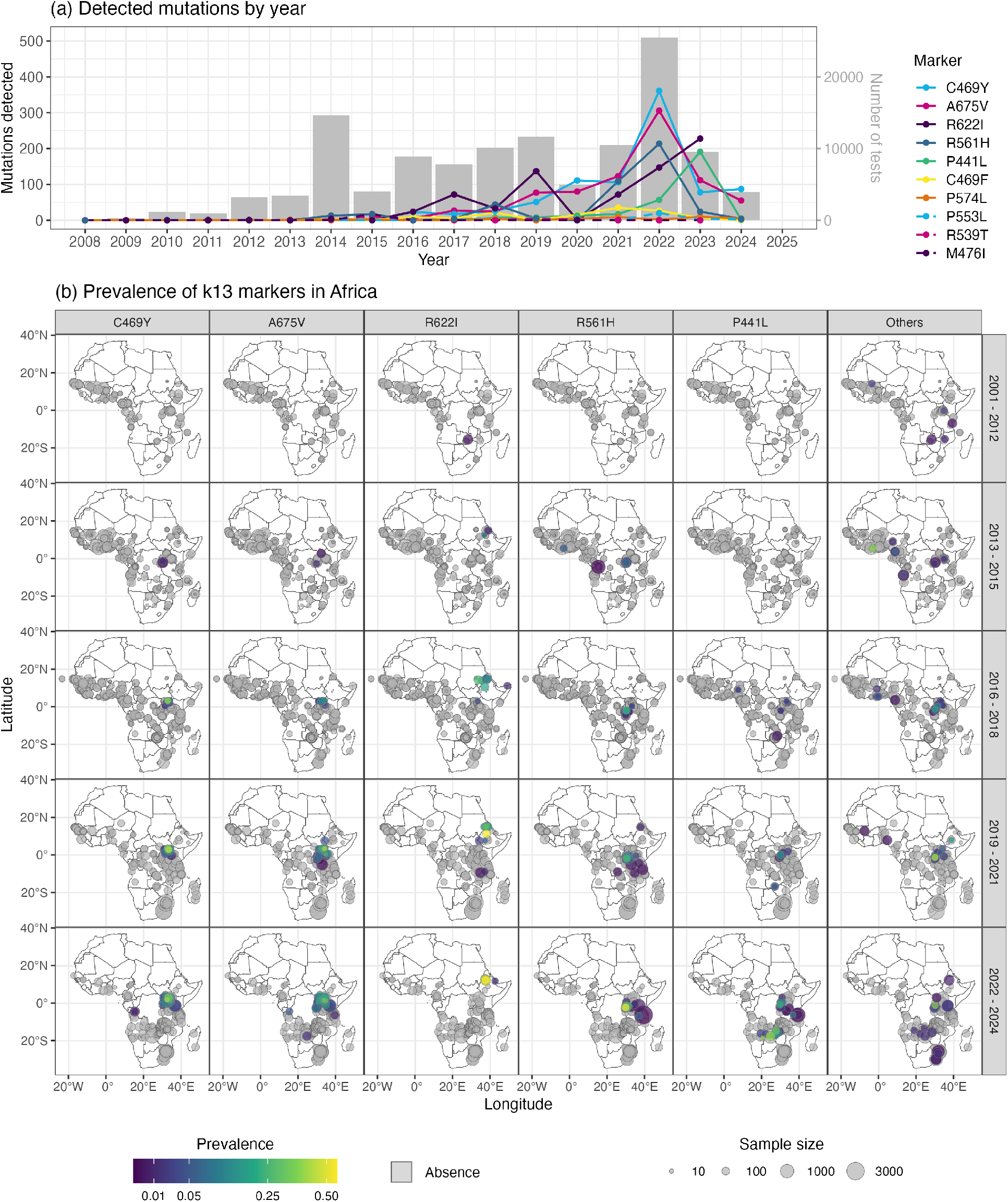
Prevalence of individual mutations in Kelch 13 dataset: (a) number of malaria cases with each mutation, 2005–2024, with total number of samples sequenced per year indicated as grey bars against the vertical axis on the right; (b) spatial distribution of the five most prevalent markers (left to right: C469Y, A675V, R622I, R561H, and P441L) over time, with all other identified mutations visualised in the right-most set of panels (each coloured point indicates aggregate prevalence of remaining ART-R-associated mutations). Point sizes are relative to the number of tests at each site. Grey points indicate surveillance detected Kelch 13 wild-types only: in other words, an absence of any of the ART-R validated or associated mutations in Table S1. All other points indicate presence of ART-R-associated Kelch 13 mutants, ranging in prevalence from 0.00081 to 0.55.

The model of aggregate Kelch 13 mutation prevalence identified all existing clusters of Kelch 13 mutations, including in the Great Lakes region and in the Horn of Africa (Figure 2). In our *nowcast* to 2026, aggregate mutation prevalence was estimated to be greater than 10% throughout Uganda, Rwanda, north-western Tanzania, north-western Kenya, north-eastern Democratic Republic of Congo (DRC), Eritrea, Ethiopia, Somalia, Sudan, and South Sudan (Figure S2). This region constitutes 23% of the area of endemic malaria transmission in Africa. Model uncertainty was highest (up to 0.12 in 2026) in areas where estimated mutation prevalence was elevated (Figure 2). Residual errors were also highest in these areas, including western Ethiopia, Uganda, and Rwanda post-2016 (Figure S3(a)), although model uncertainty *relative* to predicted prevalence was low in these regions (Figure S4). Unscaled model uncertainty was greatest in areas of low estimated prevalence and low surveillance intensity, including in Western and Central Africa (Figure S5).

**Figure 2.**
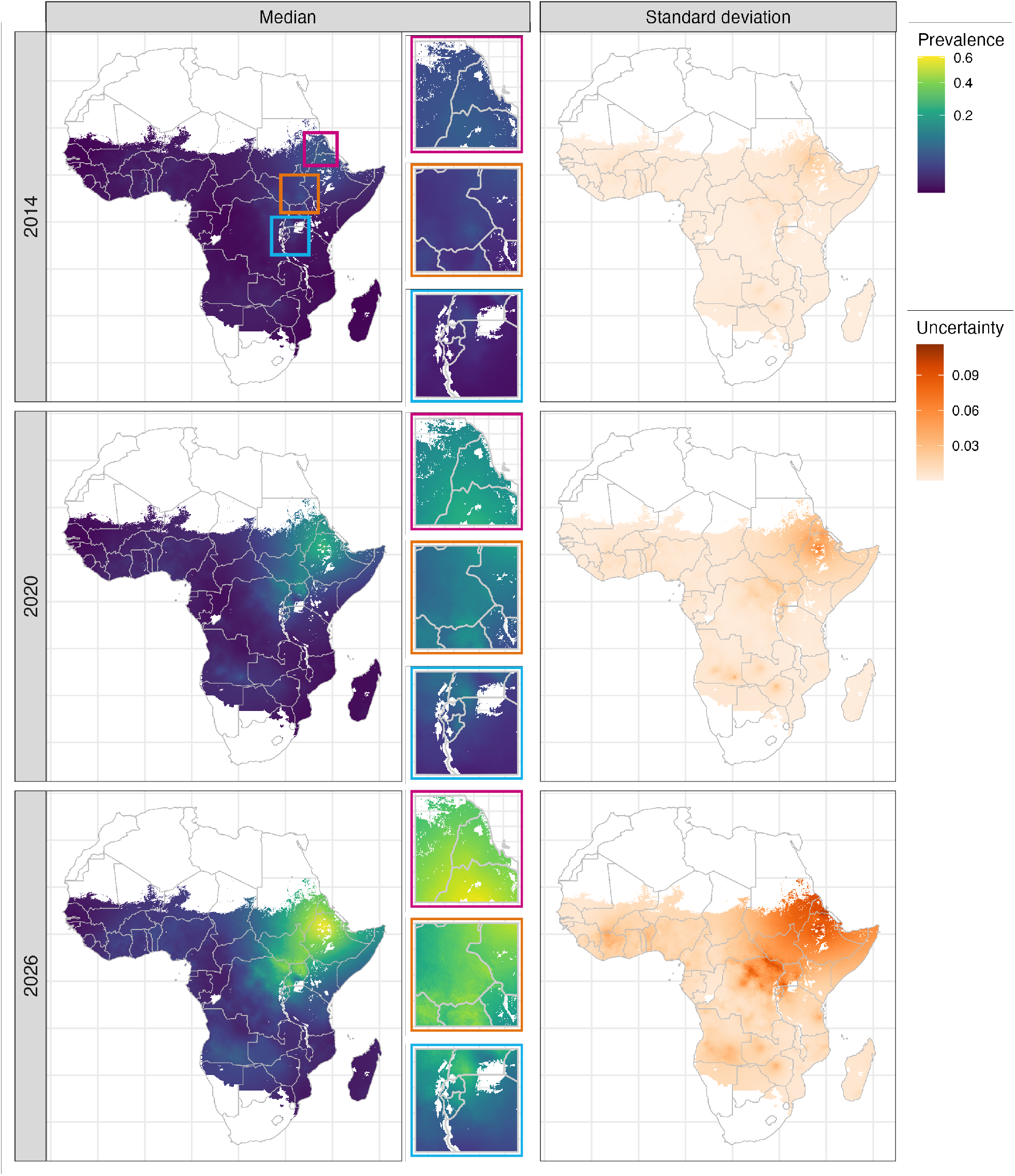
Median (left) and standard deviation (right) estimated aggregate Kelch 13 mutation prevalence in 2014, 2020, and 2026. Three regions are highlighted: the area bordering Eritrea, Ethiopia, and Sudan (magenta), northern Uganda, South Sudan, north-eastern DRC (orange), and the Great Lakes region, including southern Uganda, Rwanda, north-western Tanzania, and Burundi (light blue). The median estimated aggregate Kelch 13 prevalence is shown in these regions in central panels. Estimates for 2026 are forward predictions: the most recent records in the surveillance dataset are from 2024.

Our individual Kelch 13 mutation models reflected local spread of a number of the mutations contributing to the continental trend of increased ART-R-associated Kelch 13 mutation prevalence (Figure 3). The Horn of Africa cluster of Kelch 13 mutations in our aggregate model (Figure 2) largely reflects detections of the validated mutation, R622I: we estimated that this mutation emerged before 2014 in north-western Ethiopia and Eritrea, in line with contemporary detections [13, 14, 43]. We estimated that the validated mutations A675V and C469Y emerged before 2016 in Uganda, in line with surveillance results [15]; model predictions also reflect later detections of both mutations in the neighbouring countries of the DRC and South Sudan [44], and Kenya [45]. We estimated that the validated mutation R561H emerged in Rwanda before 2016, in line with recorded detections [46]; our model indicates subsequent spread to Tanzania, as observed by [47] and [48]. The candidate mutation P441L has been reported at prevalences greater than 30% in Southern Africa including in the Zambezi region of north-eastern Namibia [36] and central Zambia [37] (detected in 81/202 and 32/103 tests at single sites, respectively); our model of P441L estimates prevalences in this region up to 12% in 2026. As with the Kelch 13 aggregate model, residual error of each of the individual Kelch 13 mutation models is greatest in areas where observed mutation prevalences are non-zero (Figures S3 and S6).

**Figure 3.**
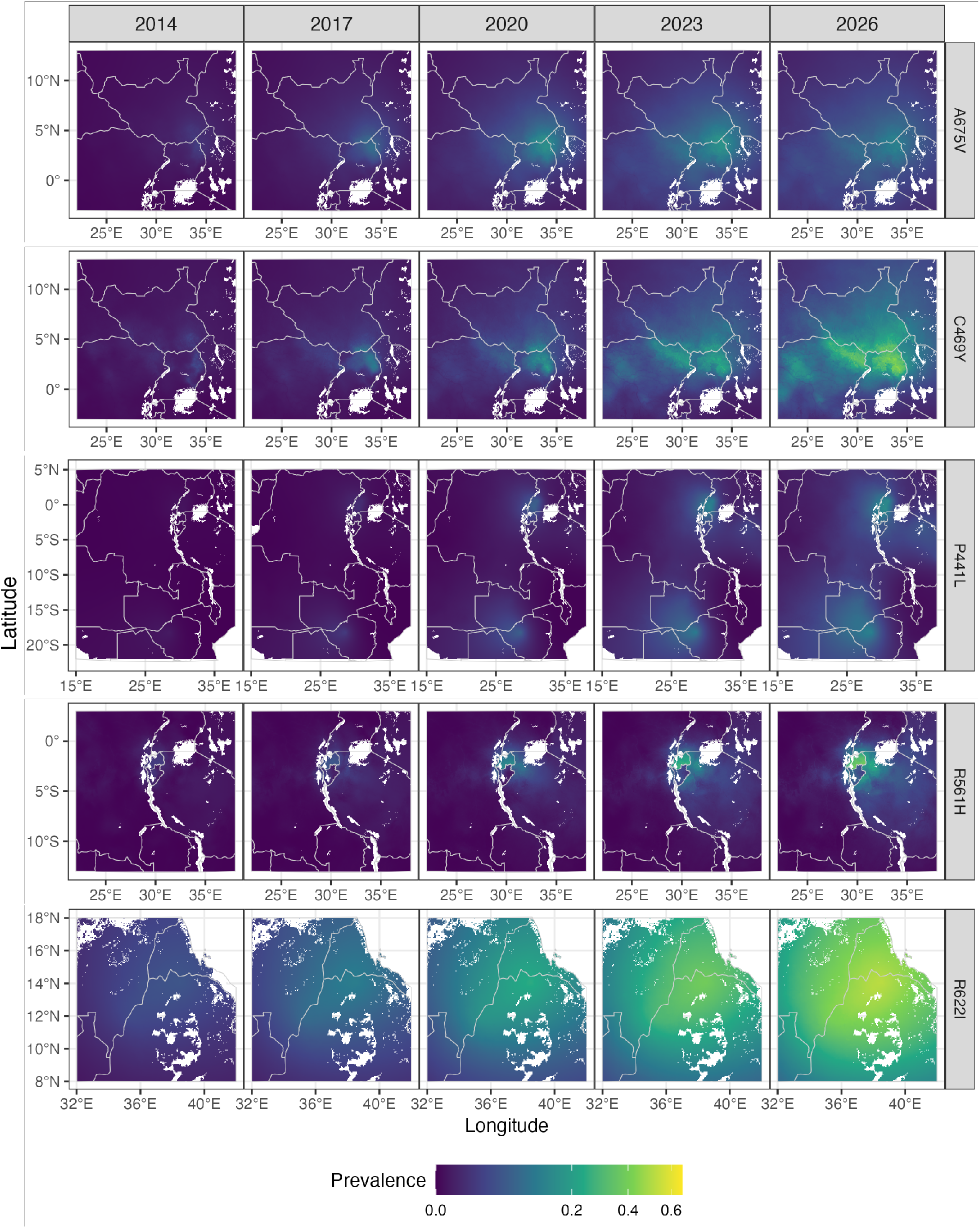
Predicted prevalence of the five most frequently detected ART-R validated or candidate Kelch 13 mutations over time, including A675V (in northern Uganda and surrounds), C469Y (in northern Uganda and surrounds), P441L (in the Zambezi region, southern Uganda, and Rwanda), R651H (in Rwanda and north-western Tanzania), and R622I (in northern Ethiopia and Eritrea). Estimates for 2026 are forward predictions: the most recent records in the surveillance dataset are from 2024.

### 3.2 Partner drug markers

Prevalences of *Pfcrt*-76T and alleles in the *Pfmdr1*-YYY haplotype, associated with reduced parasite susceptibility to amodiaquine, are estimated to decrease over the study period, and be replaced by *Pfcrt*-K76 and alleles in the *Pfmdr1*-NFD haplotype, associated with reduced parasite susceptibility to lumefantrine (Figures 4, 5, 6 and S7). This transition in marker prevalences follows the transition in drug pressure from chloroquine in the later half of the 20th century to pressure from ACTs, initially predominantly AL in Eastern and Southern Africa [49]. We estimate the median prevalence of *Pfcrt*-76T across Sub-Saharan Africa to be 19% in 2026; *Pfcrt*-76T is sustained only in the Horn of Africa and to a lesser extent, in Western Africa. In our model of *Pfmdr1*-N86Y, the wild-type allele *Pfmdr1*-N86 is estimated to be dominant throughout the area of endemic malaria transmission by as early as 2014. Our estimates of *Pfmdr1*-Y184F prevalence reflect relatively higher prevalences of the wild-type allele *Pfmdr1*-Y184 in Southern and Western Africa, including western Nigeria, in contrast to relatively higher prevalences of the mutant allele *Pfrmdr1*-184F in North-eastern Africa. *Pfmdr1*-1246Y was prevalent in Eastern Africa in 2002, but by 2020, *Pfmdr1*-D1246 is estimated to be the dominant allele throughout the area of endemic malaria transmission. Unscaled uncertainties were relatively high for the *Pfmdr1*-D1246Y model (Figures S8), which resulted from relatively high variation in spatiotemporally proximal records, for example in Uganda where the same sites were surveyed in adjacent years, identified in our assessment of residual errors (Figure S9(d)).

**Figure 4.**
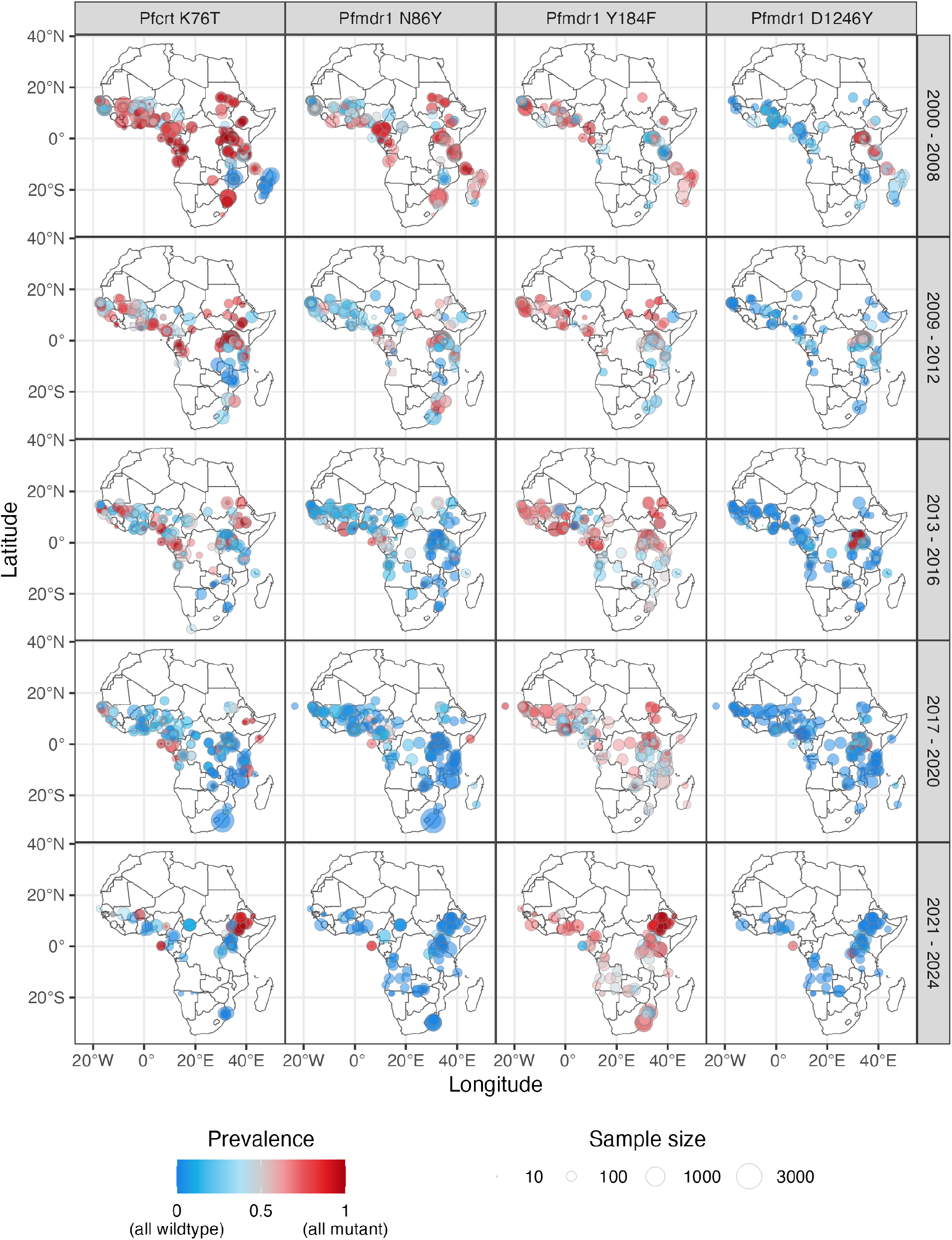
Data of surveillance of *Pfcrt*-K76T, *Pfmdr1*-N86Y, *Pfmdr1*-Y184F, and *Pfmdr1*-D1246Y, 2000– 2024. Point sizes indicate number of tests at a site. Opacity is equal for all points.

**Figure 5.**
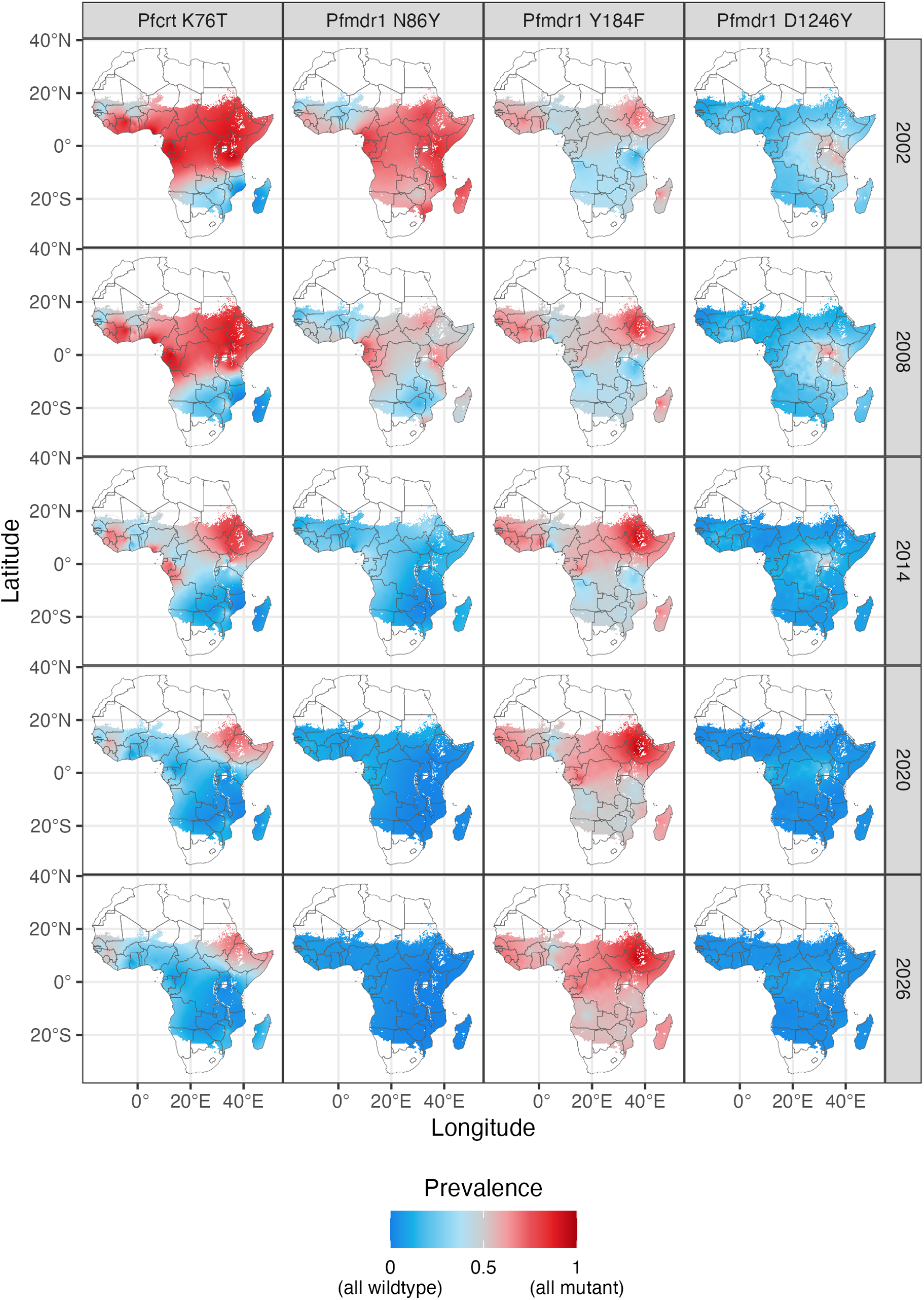
Median predicted prevalence of molecular markers associated with reduced ACT partner drug susceptibility: *Pfcrt*-K76T, *Pfmdr1*-N86Y, *Pfmdr1*-Y184F, and *Pfmdr1*-D1246Y. See corresponding uncertainty maps in Figures S7 and S8.

**Figure 6.**
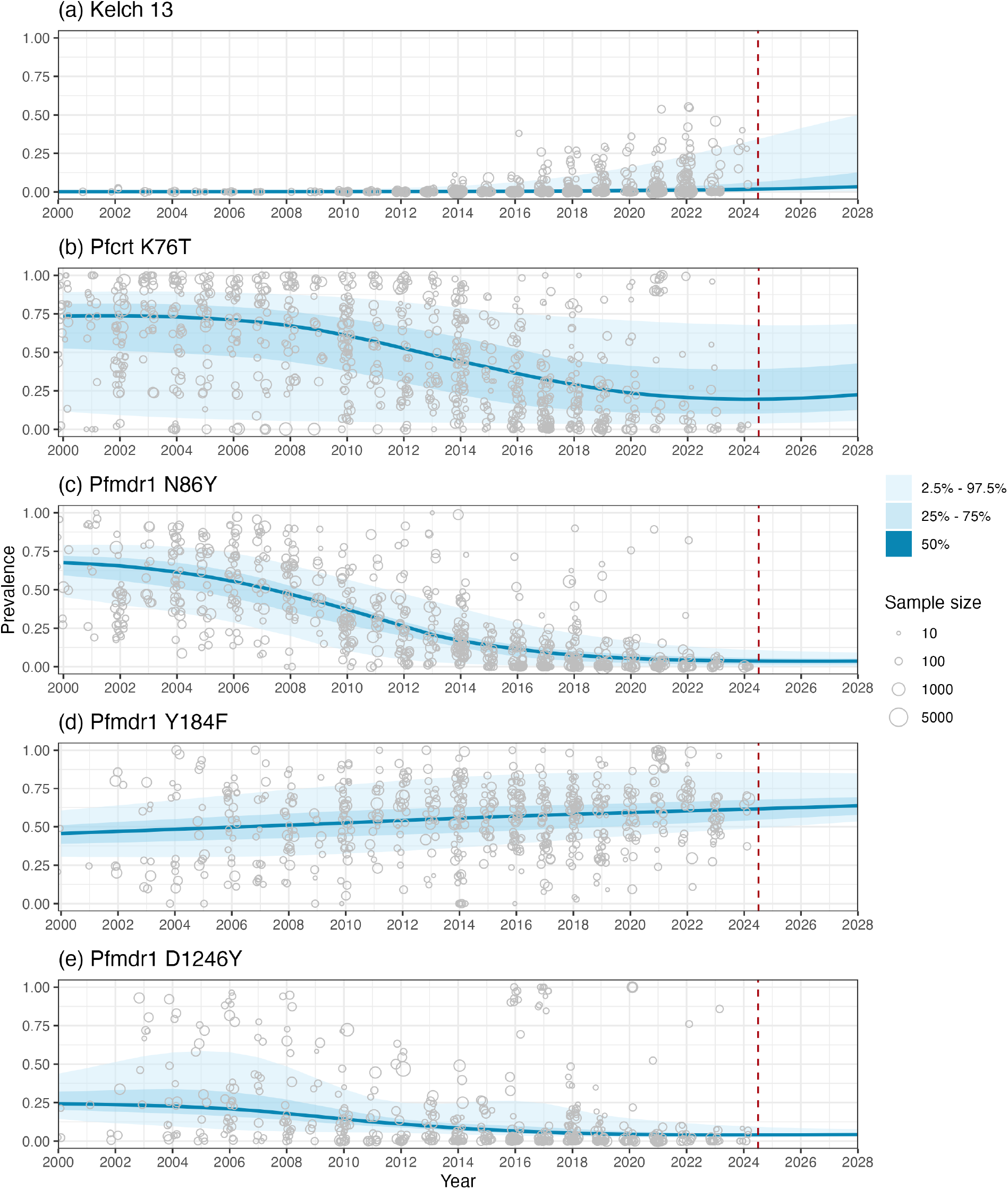
Summary of median predicted prevalence of (a) all ART-R-associated Kelch 13 mutations (b) *Pfcrt*-K76T, (c) *Pfmdr1*-N86Y, (d) *Pfmdr1*-Y184F, and (e) *Pfmdr1*-D1246Y over time. Quantiles are of all median predicted prevalences in the area of endemic transmission of *P. falciparum* malaria in Africa. Prevalence data are overlaid: the most recent sample in the prevalence dataset was collected in 2024 (vertical dashed red line indicates predictions beyond this period).

### 3.3 Trends over time

We estimate that by 2026, 5.9% (95% credible interval [3.0%, 11.1%]) of annual *P. falciparum* malaria cases in Sub-Saharan Africa will involve Kelch 13-mutant parasites associated with ART-R (Figure 7). Incidence-adjusted estimates of Kelch 13 mutation prevalence (see Supplementary Methods Section S3) include high numbers of Kelch 13-mutant infections where unadjusted estimated prevalence of Kelch 13 mutations is low but malaria incidence is high, for example in Nigeria, where approximately 24% of global malaria cases occur [1] (and where ART-R-associated Kelch 13 mutations have been only infrequently detected [50, 51]).

**Figure 7.**
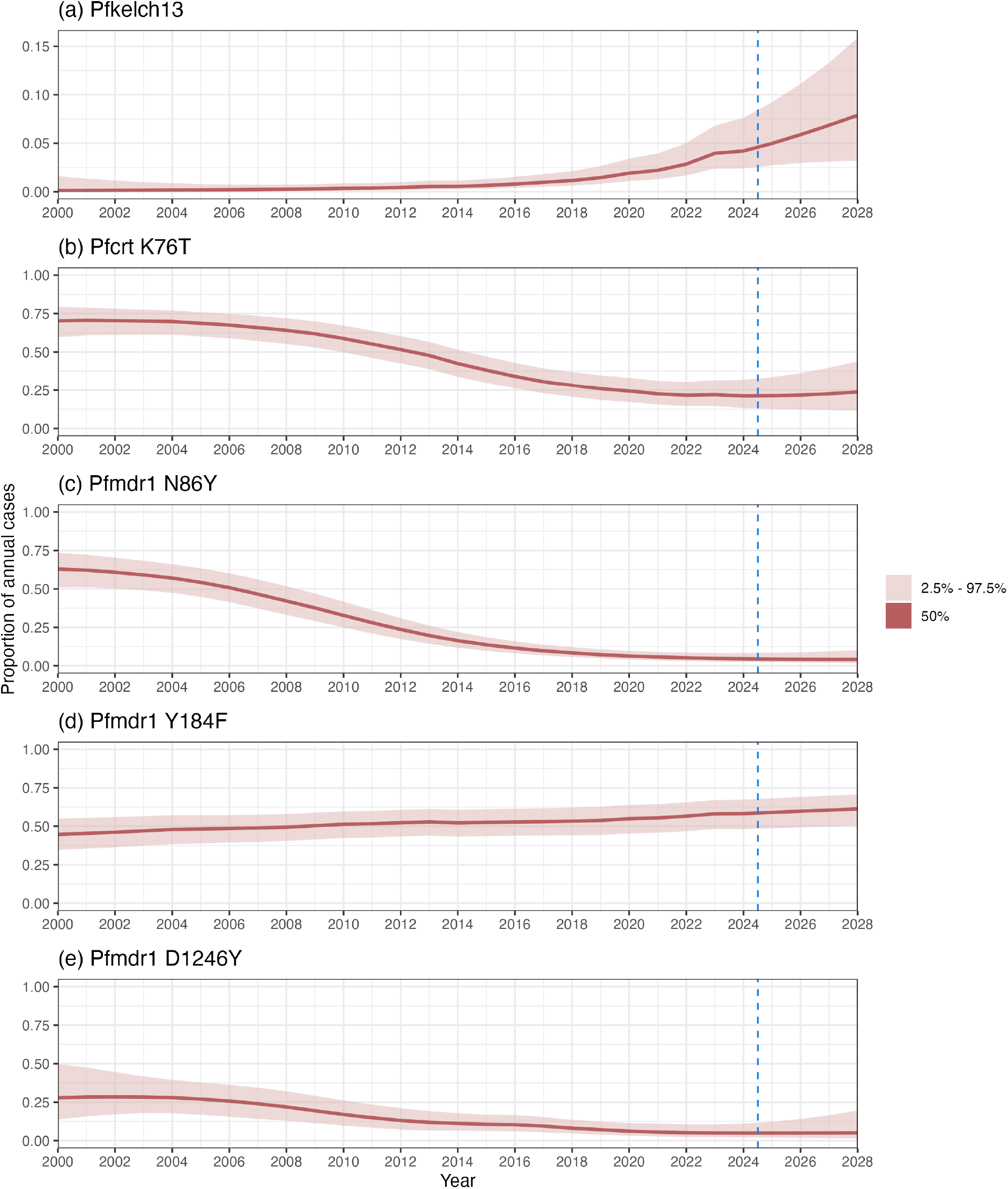
Estimated proportion of malaria cases with (a) all ART-R-associated Kelch 13 mutations (b) *Pfcrt*-K76T, (c) *Pfmdr1*-N86Y, (d) *Pfmdr1*-Y184F, and (e) *Pfmdr1*-D1246Y over time. Estimates were calculated by multiplying the Malaria Atlas Project’s gridded annual point estimates of malaria incidence [42] by the median estimate of marker prevalence, together with surfaces of the upper and lower bounds of the 95% credible interval of estimated marker prevalence. Each of the resulting three surfaces was then summed and divided by the sum of the corresponding annual incidence surface. The vertical axis range is different across the panels. (The most recent record in the marker prevalence dataset was collected in 2024: the vertical dashed blue line indicates marker prevalence predictions beyond this period.)

### 3.4 Model validation

We validate our models by assessing coverage probabilities and carrying out posterior predictive checks (see Supplementary Methods Section S2.3 for details). Model coverage probabilities, passed through the beta-binomial observation model, are inflated by the proportion of zero-prevalence records in the surveillance dataset, particularly for the Kelch 13 model (Figure S10). Probability integral transform empirical cumulative distribution functions (PIT-ECDFs) were near-uniform for partner drug marker models, indicating good fit, although the high proportion of zeroes in the Kelch 13 dataset skewed the Kelch 13 models’ PIT-ECDFs (Figure S10(c)). Residual errors were highest at sites where there was variation in spatiotemporally proximal surveillance (Figures S3, S6, and S9). Modelled estimates were all an improvement on baseline estimates, and models trained on held-out datasets performed similarly to models fitted to the full surveillance dataset (Table S2).

## 4 Discussion

We describe a spatiotemporal model of aggregated Kelch 13 mutation prevalence in *P. falciparum* malaria, as proxy for the prevalence of ART-R in Sub-Saharan Africa. To further support operational decision-making, we also predict individual prevalences of each of the five most frequently reported ART-R-associated Kelch 13 mutations in Africa, and predict prevalences of putative markers of reduced susceptibility to the most widely-used partner drugs in Africa, lumefantrine and amodiaquine, at the loci *Pfcrt*-76, *Pfmdr1*-86, *Pfmdr1*-184, and *Pfmdr1*-1246.

We estimate that Kelch 13 mutation prevalence in Africa is elevated in at least two clusters: in the Great Lakes region including Uganda, Rwanda, and Tanzania, and in North-eastern Africa including Ethiopia and Eritrea. The prevalence of markers for reduced parasite susceptibility to amodiaquine, *Pfcrt*-76T and alleles in the haplotype *Pfmdr1*-YYY has decreased during the last two decades: *Pfcrt*-K76 and alleles in the haplotype *Pfmdr1*-NFD are now the most prevalent alleles at these loci. Collectively, these alleles are markers of reduced parasite susceptibility to lumefantrine, the partner drug in AL, by far the most widely-procured ACT in Africa [1]. An exception is the relatively high estimated prevalence of *Pfcrt*-76T in Ethiopia, where *P. vivax* co-circulates and the national policy for *P. vivax* treatment is chloroquine in combination with primaquine, and to a lesser extent, in West Africa where ASAQ is still used [1].

Of note is the recent emergence of Kelch 13 mutations in the Zambezi region of Namibia and neighbouring Zambia, in particular the candidate ART-R marker P441L [1]: we estimate prevalence of Kelch 13 mutations in southern Zambia to be greater than 3% in 2026 and increasing (Figure S2). The ART-R validated mutation P574L has also recently been detected in South Africa [52]. In light of recent increases in malaria case numbers in Southern Africa [53], it is critical that drug resistance marker prevalence and ACT therapeutic efficacy in the region is monitored for changes [36].

In this work, we draw on the most comprehensive dataset of genomic surveillance of *P. falciparum* available to date, which includes both published and unpublished records. The lag time between sample collection and publication of records in the WWARN Molecular Surveyor dataset has previously been found to have a median of three years [34, 54]: it is likely that many samples collected in recent years are not yet in the dataset. Unpublished data have been provided to platforms including the MARC SE-Africa antimalarial resistance dashboard [5], and the WHO Malaria Threats Map [27], to enable more timely analysis by other researchers. It is critical that efforts to expedite the analysis and sharing of molecular surveillance of *P. falciparum* are maintained to support regional antimalarial resistance monitoring into the future.

Geostatistical modelling has the potential to add considerable value to existing molecular surveillance data. By making predictions to the entire region of endemic malaria transmission in Sub-Saharan Africa, we are able to estimate the prevalence of drug resistance markers in areas that have not been subject to surveillance. Our models are able to *nowcast* —to predict recent or current marker prevalence before the results of recent surveillance are available —and to forecast marker prevalences into the future. Our models are also associated with comprehensive estimates of prediction uncertainty: by considering both model predictions and uncertainty, decision-makers will be supported to make robust decisions.

The recent models described by Young et al. [24], based on a similar molecular surveillance dataset, produced predictions with several key differences to ours, including sustained prevalences of *Pfcrt-76T* and *Pfmdr1-86Y* in Central Africa and *Pfmdr1-86Y* in North-eastern Africa. The distinct predictions from our models and those described by Young et al. demonstrate the sensitivity of geostatistical models to methodological choices. This highlights the importance of placing models in the context of uncertainty and carrying out formal model validation, which is omitted by Young et al., so that decision-makers can be best informed about the limitations of model predictions.

While we present our models of mutation prevalence as proxies for the spatiotemporal distribution of parasite sensitivity to artemisinin, lumefantrine, amodiaquine, and other antimalarials, parasite genetics are not perfect predictors of treatment efficacy in patients. A future improvement to the models described here would be to predict a net surface of *ACT susceptibility* through a hierarchical linking of molecular sequencing and clinical parasite clearance data [55]. The effect of each of the target markers on clinical outcome may not be perfectly additive: understanding the effect of haplotypes on treatment failure rates, and creating models of haplotype prevalence (e.g., [35]), is another possible future direction. We simplify Kelch 13 data from a series of mutations to a single, aggregated dataset, under the assumption that each of the mutations highlighted in Table S1 are of equivalent interest to decision-makers. However, we acknowledge that each of these mutations has distinct spatiotemporal distributions (Figure 1), and may have variable effects on parasite susceptibility to artemisinin [55]. It is for this reason that we also describe models of the five ART-R-associated Kelch 13 mutations most frequently identified in Africa.

There are several possible policy changes that can be made to respond to rising ART-R: the use of multiple first-line treatments and the use of triple ACTs (an artemsinin derivative in combination with *two* partner drugs, such as lumefantrine and amodiaquine), make adaptive use of existing antimalarials and exploit opposing selective pressures [56]. Prophylactic chemoprevention strategies, in children and pregnant women, have the potential to protect vulnerable groups within the population, but may facilitate the evolution of drug resistance through increased parasite exposure to ACT partner drugs such as amodiaquine [57]. It is critical that the deployment of each of these control measures is guided by accurate, precise estimates of the current and future predictive distributions of antimalarial drug resistance. In the past, implementing malaria treatment guidelines has taken 18 to 32 months (e.g., [58]); malaria treatment policy changes cannot be driven while looking in the rear view mirror and policy-makers must be able to see the road ahead. Geospatial models drawing on molecular surveillance data have the potential to provide validated nowcast and forecast estimates of mutation prevalences, contextualised by model uncertainty, to guide operational decision-making.

Our model of aggregate Kelch 13 mutation prevalence summarises existing genomic surveillance data and makes predictions at locations and times where data were not collected, or have not yet been published. Geospatial models have the potential to guide researchers and national malaria programme decisions on where sample collections will be most cost-effective and informative. Interpreted together, our models build a picture of the spatiotemporal distribution of parasite sensitivity to the artemisinin combinations AL and ASAQ, the most widely-used first-line treatments for uncomplicated malaria in Africa. Our models have the potential to guide the collection of surveillance data and the deployment of control measures into the future. In the current global health funding climate, and in the face of recent stagnation in progress towards malaria elimination in Africa, it is critical that decision and policy-making is quantitatively guided by data and validated modelling.

## Data Availability

All data analysed in the present study are available online at https://surveyor.iddo.org and https://github.com/Stephanie-van-Wyk/MARC_SEA_dashboard. All code for analysis is available online at https://github.com/Infectious-Diseases-Data-Observatory/act_markers_maps.

## 5 Acknowledgements

This research was funded by the European Union under the Global Health EDCTP3 Joint Undertaking (grant agreement 101103076) as part of the Mitigating Antimalarial Resistance Consortium in South-East Africa (MARC SE-Africa) and the Australian National Health and Medical Research Council (APP2019093). J.A. Flegg’s research is supported by the Australian Research Council (FT210100034, CE230100001).

## References

[1] World Health Organization. World malaria report 2025. World Health Organization, 2025.

[2] Nicholas J White, Tran T Hien, and François H Nosten. “A brief history of Qinghaosu”. Trends in parasitology 31:12 (2015), pp. 607–610. doi: 10.1016/j.pt.2015.10.010.

[3] François Nosten, Dominique Richard-Lenoble, and Martin Danis. “A brief history of malaria”. La presse médicale 51:3 (2022), p. 104130. doi: 10.1016/j.lpm.2022.104130.

[4] Jennifer A Flegg, Charlotte JE Metcalf, Myriam Gharbi, Meera Venkatesan, Tanya Shewchuk, Carol Hopkins Sibley, and Philippe J Guerin. “Trends in antimalarial drug use in Africa”. The American Journal of Tropical Medicine and Hygiene 89:5 (2013), p. 857. doi: 10.4269/ajtmh.13-0129.

[5] Stephanie van Wyk, Ishen Seocharan, Eulambius M Mlugu, Dhol S Ayuen, Donnie Mategula, Tikhala Makhaza, James Kiarie, Victor Asua, Jimmy Opigo, Aimable Mbituyumuremyi, et al. “The MARC SE-Africa Dashboard: Joining forces to counteract emerging antimalarial resistance in south and east Africa”. medRxiv (2025), pp. 2025–01. doi: 10.1101/2025.01.07.25320158.

[6] Giulia Rathmes, Susan F. Rumisha, Tim C. D. Lucas, Katherine A. Twohig, Andre Python, Michele Nguyen, Anita K. Nandi, Suzanne H. Keddie, Emma L. Collins, Jennifer A. Rozier, et al. “Global estimation of anti-malarial drug effectiveness for the treatment of uncomplicated Plasmodium falciparum malaria 1991–2019”. Malaria Journal 19:1 (2020), p. 374. doi: 10.1186/s12936-020-03446-8.

[7] Harald Noedl, Youry Se, Kurt Schaecher, Bryan L Smith, Duong Socheat, and Mark M Fukuda. “Evidence of artemisinin-resistant malaria in western Cambodia”. New England Journal of Medicine 359:24 (2008), pp. 2619–2620. doi: 10.1056/NEJMc080501.

[8] Arjen M Dondorp, François Nosten, Poravuth Yi, Debashish Das, Aung Phae Phyo, Joel Tarning, Khin Maung Lwin, Frederic Ariey, Warunee Hanpithakpong, Sue J Lee, et al. “Artemisinin resistance in Plasmodium falciparum malaria”. New England journal of medicine 361:5 (2009), pp. 455–467. doi: 10.1056/NEJMoa0808859.

[9] Elizabeth A Ashley, Mehul Dhorda, Rick M Fairhurst, Chanaki Amaratunga, Parath Lim, Seila Suon, Sokunthea Sreng, Jennifer M Anderson, Sivanna Mao, Baramey Sam, et al. “Spread of artemisinin resistance in Plasmodium falciparum malaria”. New England Journal of Medicine 371:5 (2014), pp. 411– 423. doi: 10.1056/NEJMoa1314981.

[10] World Health Organization. Compendium of molecular markers for antimalarial drug resistance. 2025. url: https://www.who.int/tools/compendium-of-molecular-markers-for-antimalarial-drug-resistance (visited on 02/24/2026).

[11] Jennifer A Flegg, Sevvandi Kandanaarachchi, Philippe J Guerin, Arjen M Dondorp, Francois H Nosten, Sabina Dahlström Otienoburu, and Nick Golding. “Spatio-temporal spread of artemisinin resistance in Southeast Asia”. PLOS Computational Biology 20:4 (2024), e1012017. doi: 10.1371/journal.pcbi.1012017.

[12] Varanya Wasakul, Tess D Verschuuren, Nguyen Thuy-Nhien, Ethan Booth, Huynh Hong Quang, Ngo Duc Thang, Keobouphaphone Chindavongsa, Siv Sovannaroth, Virasak Banouvong, Viengphone Sengsavath, et al. “Genetic surveillance of Plasmodium falciparum populations following treatment policy revisions in the Greater Mekong Subregion”. Nature Communications 16:1 (2025), p. 4689. doi: 10.1038/s41467-025-59946-1.

[13] Abebe Genetu Bayih, Gebeyaw Getnet, Abebe Alemu, Sisay Getie, Abu Naser Mohon, and Dylan R Pillai. “A unique Plasmodium falciparum Kelch 13 gene mutation in northwest Ethiopia”. The American journal of tropical medicine and hygiene 94:1 (2016), p. 132. doi: 10.4269/ajtmh.15-0477.

[14] Mariangela L’Episcopia, Julia Kelley, Dhruviben Patel, Sarah Schmedes, Shashidahar Ravishankar, Michela Menegon, Edvige Perrotti, Abduselam M Nurahmed, Albadawi A Talha, Bakri Y Nour, et al. “Targeted deep amplicon sequencing of kelch 13 and cytochrome b in Plasmodium falciparum isolates from an endemic African country using the Malaria Resistance Surveillance (MaRS) protocol”. Parasites & Vectors 13:1 (2020), p. 137. doi: 10.1016/j.ijid.2021.04.081.

[15] Costanza Tacoli, Prabhanjan P Gai, Claude Bayingana, Kevin Sifft, Dominik Geus, Jules Ndoli, Augustin Sendegeya, Jean Bosco Gahutu, and Frank P Mockenhaupt. “Artemisinin resistance–associated K13 polymorphisms of Plasmodium falciparum in Southern Rwanda, 2010–2015”. The American journal of tropical medicine and hygiene 95:5 (2016), p. 1090. doi: 10.4269/ajtmh.16-0483.

[16] Abdoulaye Djimdé, Ogobara K Doumbo, Joseph F Cortese, Kassoum Kayentao, Safi Doumbo, Yacouba Diourté, Drissa Coulibaly, Alassane Dicko, Xin-zhuan Su, Takashi Nomura, et al. “A molecular marker for chloroquine-resistant falciparum malaria”. New England journal of medicine 344:4 (2001), pp. 257– 263. doi: 10.1056/NEJM200101253440403.

[17] GS Humphreys, I Merinopoulos, J Ahmed, CJM Whitty, TK Mutabingwa, CJ Sutherland, and RL Hallett. “Amodiaquine and artemether-lumefantrine select distinct alleles of the Plasmodium falciparum mdr1 gene in Tanzanian children treated for uncomplicated malaria”. Antimicrobial agents and chemotherapy 51:3 (2007), pp. 991–997. doi: 10.1128/aac.00875-06.

[18] Gabrielle Holmgren, Johan Hamrin, Jenny Svärd, Andreas Mårtensson, José Pedro Gil, and Anders Björkman. “Selection of pfmdr1 mutations after amodiaquine monotherapy and amodiaquine plus artemisinin combination therapy in East Africa”. Infection, genetics and evolution 7:5 (2007), pp. 562– 569. doi: 10.1016/j.meegid.2007.03.005.

[19] Samuel L Nsobya, Christian Dokomajilar, Moses Joloba, Grant Dorsey, and Philip J Rosenthal. “Resistance-mediating Plasmodium falciparum pfcrt and pfmdr1 alleles after treatment with artesunate-amodiaquine in Uganda”. Antimicrobial agents and chemotherapy 51:8 (2007), pp. 3023–3025. doi: 10.1128/aac.00012-07.

[20] Meera Venkatesan, Nahla B Gadalla, Kasia Stepniewska, Prabin Dahal, Christian Nsanzabana, Clarissa Moriera, Ric N Price, Andreas Mårtensson, Philip J Rosenthal, Grant Dorsey, et al. “Polymorphisms in Plasmodium falciparum chloroquine resistance transporter and multidrug resistance 1 genes: parasite risk factors that affect treatment outcomes for P. falciparum malaria after artemether-lumefantrine and artesunate-amodiaquine”. The American journal of tropical medicine and hygiene 91:4 (2014), p. 833. doi: 10.4269/ajtmh.14-0031.

[21] Alexandra Walker, Amanda Ross, and Christian Nsanzabana. “Changes in the Prevalence of Antimalarial Partner Drug Resistance Markers and Policy in 6 Sub-Saharan African Countries From 2000 to 2021: A Systematic Review”. Open Forum Infectious Diseases. Vol. 12. 9. Oxford University Press US. 2025, ofaf508. doi: 10.1093/ofid/ofaf508.

[22] Lucy C Okell, Lisa Malene Reiter, Lene Sandø Ebbe Vito Baraka, Donal Bisanzio, Oliver J Watson, Adam Bennett, Robert Verity, Peter Gething, Cally Roper, et al. “Emerging implications of policies on malaria treatment: genetic changes in the Pfmdr-1 gene affecting susceptibility to artemether– lumefantrine and artesunate–amodiaquine in Africa”. BMJ global health 3:5 (2018). doi: 10.1136/bmjgh-2018-000999.

[23] Andrew J Balmer, Nina FD White, Eyyüb S Ünlü, Chiyun Lee, Richard D Pearson, Jacob Almagro-Garcia, and Cristina Ariani. “Understanding the global rise of artemisinin resistance: Insights from over 100,000 Plasmodium falciparum samples”. eLife 14 (2025), e105544. doi: 10.7554/eLife.105544.

[24] Neeva Wernsman Young, Cécile PG Meier-Scherling, Gina Cuomo-Dannenburg, George A Tollefson, Sean V Connelly, Jacob Marglous, Isabela Gerdes Gyuricza, Kelly Carey-Ewend, Ronald Kyong-Shin, Zachary R Popkin-Hall, et al. “Mapping the prevalence of molecular markers of Plasmodium falciparum artemisinin partial resistance in Africa: a spatial-temporal modelling study”. medRxiv (2025). doi: 10.64898/2025.12.22.25342873.

[25] Infectious Diseases Data Observatory. IDDO Molecular Surveyor. 2026. url: https://surveyor.iddo.org (visited on 02/24/2026).

[26] Sabina Dahlström Otienoburu, Ignacio Suay, Steven Garcia, Nigel V Thomas, Suttipat Srisutham, Anders Björkman, and Georgina S Humphreys. “An online mapping database of molecular markers of drug resistance in Plasmodium falciparum: the ACT Partner Drug Molecular Surveyor”. Malaria journal 18 (2019), pp. 1–10. doi: 10.1186/s12936-019-2645-x.

[27] World Health Organization. WHO Malaria Threats Map: molecular marker studies of antimalarial drug resistance. 2025. url: https://apps.who.int/malaria/maps/threats (visited on 02/24/2026).

[28] Christian Nsanzabana, Djibrine Djalle, Philippe J Guérin, Didier Ménard, and Iveth J González. “Tools for surveillance of anti-malarial drug resistance: an assessment of the current landscape”. Malaria journal 17:1 (2018), p. 75. doi: 10.1186/s12936-018-2185-9.

[29] Pierre Gashema, James Kagame, Patrick Gad Iradukunda, Emmanuel Edwar Siddig, Sofonias Kifle Tessema, Merawi Aragaw Tegegne, Mazyanga Lucy Mazaba, Mosoka Fallah, Daniel Ngamije, Jean de Dieu Harelimana, et al. “Mapping Plasmodium falciparum mutations in Africa: A critical review of emerging drug resistance and implications for malaria control.” International Journal of Infectious Diseases (2025), p. 108033. doi: 10.1016/j.ijid.2025.108033.

[30] Shazia Ruybal-Pesántez, Kirsty McCann, Jessy Vibin, Sasha Siegel, Sarah Auburn, and Alyssa E Barry. “Molecular markers for malaria genetic epidemiology: progress and pitfalls”. Trends in parasitology 40:2 (2024), pp. 147–163. doi: 10.1016/j.pt.2023.11.006.

[31] Apoorv Gupta, Lucinda E Harrison, Minu Nain, Sauman Singh Phulgenda, Rutuja Chhajed, Roopal S Kumar, Aishika Das, Manju Rahi, Philippe J Guerin, Anup R Anvikar, et al. “Model-guided geospatial surveillance system for antimalarial drug resistance”. PLOS Global Public Health 6:1 (2026), e0004717. doi: 10.1371/journal.pgph.0004717.

[32] Lucinda E Harrison, Jennifer A Flegg, Ruarai Tobin, Inke ND Lubis, Rintis Noviyanti, Matthew J Grigg, Freya M Shearer, and David J Price. “A multi-criteria framework for disease surveillance site selection: case study for Plasmodium knowlesi malaria in Indonesia”. Royal Society Open Science 11:1 (2024), p. 230641. doi: 10.1098/rsos.230641.

[33] Minu Nain, Mehul Dhorda, Jennifer A Flegg, Apoorv Gupta, Lucinda E Harrison, Sauman Singh-Phulgenda, Sabina D Otienoburu, Eli Harriss, Praveen K Bharti, Beauty Behera, et al. “Systematic Review and Geospatial Modeling of Molecular Markers of Resistance to Artemisinins and Sulfadoxine– Pyrimethamine in Plasmodium falciparum in India”. The American journal of tropical medicine and hygiene 110:5 (2024), p. 910. doi: 10.4269/ajtmh.23-0631.

[34] Jennifer A Flegg, Georgina S Humphreys, Brenda Montanez, Taryn Strickland, Zaira J Jacome-Meza, Karen I Barnes, Jaishree Raman, Philippe J Guerin, Carol Hopkins Sibley, and Sabina Dahlström Otienoburu. “Spatiotemporal spread of Plasmodium falciparum mutations for resistance to sulfadoxine-pyrimethamine across Africa, 1990–2020”. PLOS Computational Biology 18:8 (2022), e1010317. doi: 10.1371/journal.pcbi.1010317.

[35] Yong See Foo and Jennifer A Flegg. “A spatio-temporal model of multi-marker antimalarial resistance”. Journal of the Royal Society Interface 21:210 (2024), p. 20230570. doi: 10.1098/rsif.2023.0570.

[36] Lydia Eloff, Andrés Aranda-Díaz, Isobel Routledge, Amy Wesolowski, Mukosha Chisenga, Brighton Mangena, John Chimumbwa, Chadwick Sikaala, Petrina Uusiku, Stark Katokele, et al. “High prevalence of molecular markers associated with artemisinin, sulphadoxine and pyrimethamine resistance in northern Namibia”. medRxiv (2025), pp. 2025–01. doi: 10.1101/2025.01.09.25320247.

[37] Andrés Aranda-Díaz, Sydney Mwanza, Takalani I Makhanthisa, Sonja B Lauterbach, Faith De Amaral, Mukosha Chisenga, Brighton Mangena, Isobel Routledge, Blaženka Letinić, Bertha Kasonde, et al. “Plasmodium falciparum genomic surveillance reveals a diversity of kelch13 mutations in Zambia”. medRxiv (2025), pp. 2025–02. doi: 10.1101/2025.02.19.25322554.

[38] Anne C Martin, Jacob M Sadler, Alfred Simkin, Michael Musonda, Ben Katowa, Japhet Matoba, Jessica Schue, Edgar Simulundu, Jeffrey A Bailey, William J Moss, et al. “Emergence and Rising Prevalence of Artemisinin Partial Resistance Marker Kelch13 P441L in a Low Malaria Transmission Setting in Southern Zambia”. The Journal of Infectious Diseases (2025), jiaf188. doi: 10.1093/infdis/jiaf188.

[39] Sabina Dahlström Otienoburu, Philippe Guerin, Stephanie van Wyk, Caitlin Richmond, Infectious Diseases Data Observatory, Farhad Shokraneh, and Dhol S Ayuen. “Prevalence Of Plasmodium falciparum Molecular Markers Associated With Resistance To Antimalarial Drugs: A Systematic Review And Meta-Analysis”. Open Science Framework (2025). doi: 10.17605/OSF.IO/8CD3A.

[40] Nick Golding and Bethan V Purse. “Fast and flexible Bayesian species distribution modelling using Gaussian processes”. Methods in Ecology and Evolution 7:5 (2016), pp. 598–608. doi: 10.1111/2041-210X.12523.

[41] Emilio Porcu, Reinhard Furrer, and Douglas Nychka. “30 Years of space–time covariance functions”. Wiley Interdisciplinary Reviews: Computational Statistics 13:2 (2021), e1512. doi: 10.1002/wics.1512.

[42] Daniel J Weiss, Paulina A Dzianach, Adam Saddler, Jailos Lubinda, Annie Browne, Michael McPhail, Susan F Rumisha, Francesca Sanna, Yalemzewod Gelaw, Juniper B Kiss, et al. “Mapping the global prevalence, incidence, and mortality of Plasmodium falciparum and Plasmodium vivax malaria, 2000– 22: a spatial and temporal modelling study”. The Lancet 405:10483 (2025), pp. 979–990. doi: 10.1016/S0140-6736(25)00038-8.

[43] Selam Mihreteab, Lucien Platon, Araia Berhane, Barbara H Stokes, Marian Warsame, Pascal Campagne, Alexis Criscuolo, Laurence Ma, Nathalie Petiot, Cécile Doderer-Lang, et al. “Increasing prevalence of artemisinin-resistant HRP2-negative malaria in Eritrea”. New England Journal of Medicine 389:13 (2023), pp. 1191–1202. doi: 10.1056/NEJMoa2210956.

[44] Stephen Tukwasibwe, Shreeya Garg, Thomas Katairo, Victor Asua, Brian A Kagurusi, Gerald Mboowa, Rebecca Crudale, Gerald Tumusiime, Julius Businge, David Alula, et al. “Varied prevalence of antimalarial drug resistance markers in different populations of newly arrived refugees in Uganda”. The Journal of Infectious Diseases 230:2 (2024), pp. 497–504. doi: 10.1093/infdis/jiae288.

[45] Victor Osoti, Kevin Wamae, Moses M Musau, John B Magudha, Leonard Ndwiga, Paul M Gichuki, Collins Okoyo, Kiplagat Rosebella, Sammy Mahugu, Stephen Aricha, et al. “Serial cross-sectional school surveys identifies C469Y, P553L, R561H and A675V kelch 13 mutations associated with artemisinin resistance in Western Kenya”. Scientific Reports 15:1 (2025), p. 38303. doi: 10.1038/s41598-025-22286-7.

[46] Aline Uwimana, Eric Legrand, Barbara H Stokes, Jean-Louis Mangala Ndikumana, Marian Warsame, Noella Umulisa, Daniel Ngamije, Tharcisse Munyaneza, Jean-Baptiste Mazarati, Kaendi Munguti, et al. “Emergence and clonal expansion of in vitro artemisinin-resistant Plasmodium falciparum kelch13 R561H mutant parasites in Rwanda”. Nature medicine 26:10 (2020), pp. 1602–1608. doi: 10.1038/s41591-020-1005-2.

[47] Jonathan J Juliano, David J Giesbrecht, Alfred Simkin, Abebe A Fola, Beatus M Lyimo, Dativa Pereus, Catherine Bakari, Rashid A Madebe, Misago D Seth, Celine I Mandara, et al. “Country wide surveillance reveals prevalent artemisinin partial resistance mutations with evidence for multiple origins and expansion of high level sulfadoxine-pyrimethamine resistance mutations in northwest Tanzania”. MedRxiv (2023). doi: 10.1101/2023.11.07.23298207.

[48] Deus S Ishengoma, Celine I Mandara, Catherine Bakari, Abebe A Fola, Rashid A Madebe, Misago D Seth, Filbert Francis, Creyton C Buguzi, Ramadhan Moshi, Issa Garimo, et al. “Evidence of artemisinin partial resistance in northwestern Tanzania: clinical and molecular markers of resistance”. The Lancet Infectious Diseases 24:11 (2024), pp. 1225–1233. doi: 10.1016/S1473-3099(24)00362-1.

[49] World Health Organization. World malaria report 2017. World Health Organization, 2017.

[50] Monday Tola, Olumide Ajibola, Emmanuel Taiwo Idowu, Olusesan Omidiji, Samson Taiwo Awolola, and Alfred Amambua-Ngwa. “Molecular detection of drug resistant polymorphisms in Plasmodium falciparum isolates from Southwest, Nigeria”. BMC research notes 13:1 (2020), p. 497. doi: 10.1186/s13104-020-05334-5.

[51] Alexandra Martín Ramírez, Akeem Abiodun Akindele, Vicenta González Mora, Luz García, Nicole Lara, Eva de la Torre-Capitán Matías, Irene Molina de la Fuente, Sulaiman Adebayo Nassar, Thuy-Huong Ta-Tang, Agustín Benito, et al. “Mutational profile of pfdhfr, pfdhps, pfmdr1, pfcrt and pfk13 genes of P. falciparum associated with resistance to different antimalarial drugs in Osun state, south-western Nigeria”. Tropical Medicine and Health 53:1 (2025), pp. 1–15. doi: 10.1186/s41182-025-00732-6.

[52] Jaishree Raman, Maxwell Mabona, Qedusizi Nyawo, Brighton Mangena, Gerdalize Kok, Lihle Mathaba, Gillian Malatje, Sonja B Lauterbach, Takalani I Makhanthisa, Hazel Gwarinda, et al. “Very Low Prevalence of Validated kelch13 Mutations and Absence of Histidine-Rich Protein 2/3 Double Gene Deletions in South African Malaria-Eliminating Districts (2022–2024)”. The American Journal of Tropical Medicine and Hygiene 1:aop (2025). doi: 10.4269/ajtmh.25-0204.

[53] Africa Centres for Disease Control and Prevention. Malaria Surge in Southern Africa. 2025. url: https://africacdc.org/news-item/malaria-surge-in-southern-africa/ (visited on 02/24/2026).

[54] Frank M Kagoro, Karen I Barnes, Kevin Marsh, Nattwut Ekapirat, Chris Erwin G Mercado, Ipsita Sinha, Georgina Humphreys, Mehul Dhorda, Philippe J Guerin, and Richard J Maude. “Mapping genetic markers of artemisinin resistance in Plasmodium falciparum malaria in Asia: a systematic review and spatiotemporal analysis”. The Lancet Microbe 3:3 (2022), e184–e192. doi: 10.1016/S2666-5247(21)00249-4.

[55] WWARN K13 Genotype-Phenotype Study Group. “Association of mutations in the Plasmodium falciparum Kelch13 gene (Pf3D7 1343700) with parasite clearance rates after artemisinin-based treatments—a WWARN individual patient data meta-analysis”. BMC medicine 17 (2019), pp. 1–20. doi: 10.1186/s12916-018-1207-3.

[56] Maciej F Boni, Nicholas J White, and J Kevin Baird. “The community as the patient in malaria-endemic areas: preempting drug resistance with multiple first-line therapies”. PLoS medicine 13:3 (2016), e1001984.

[57] Christopher V Plowe. “Malaria chemoprevention and drug resistance: a review of the literature and policy implications”. Malaria Journal 21:1 (2022), p. 104. doi: 10.1186/s12936-022-04115-8.

[58] Naawa Sipilanyambe, Jonathon L Simon, Pascalina Chanda, Peter Olumese, Robert W Snow, and Davidson H Hamer. “From chloroquine to artemether-lumefantrine: the process of drug policy change in Zambia”. Malaria Journal 7:1 (2008), p. 25. doi: 10.1186/1475-2875-7-25.

