## Supplementary figures for "Estimating the changing prevalence of molecular markers of artemisinin partial resistance in *Plasmodium falciparum* malaria in Sub-Saharan Africa"

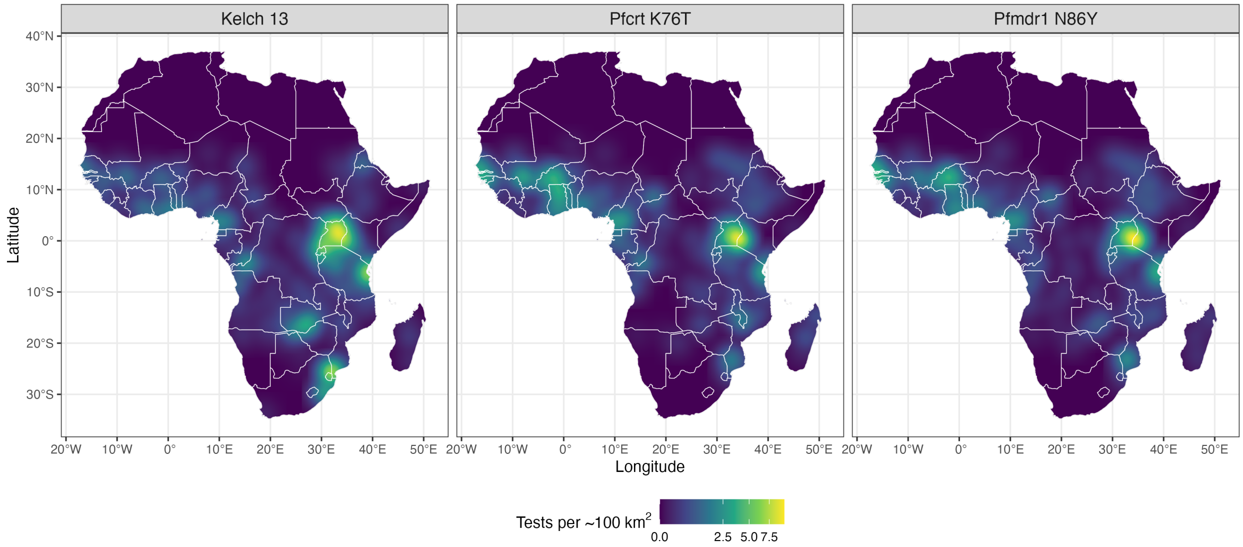


Figure S1: Surveillance intensity for Kelch 13, *Pfcrt*-K76T, and *Pfmdr1*-N86Y: Gaussian kernel density estimate on the number of tests in a pixel, with standard deviation equal to 1.5°. Units of $km^2$ are approximate, using the conversion 1°= 111 km. Of the *Pfmdr1* markers modelled in this manuscript, only *Pfmdr1*-N86Y is shown here for brevity: surveillance effort was roughly equivalent for all three *Pfmdr1* markers.

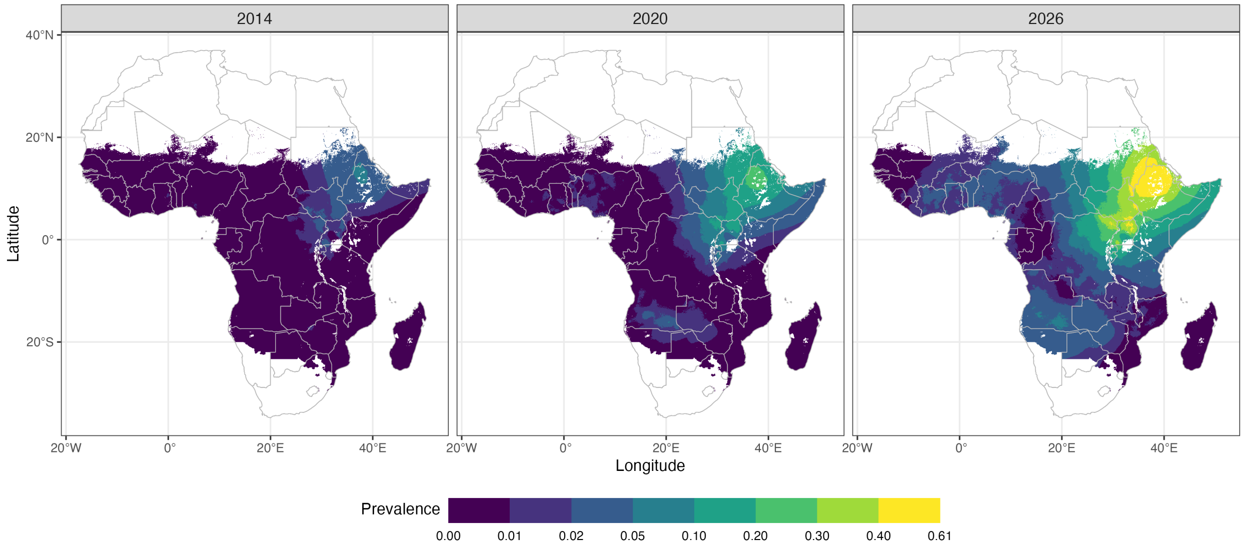


Figure S2: Contours of median predicted aggregate Kelch 13 mutation prevalence in 2014, 2020, and 2026.

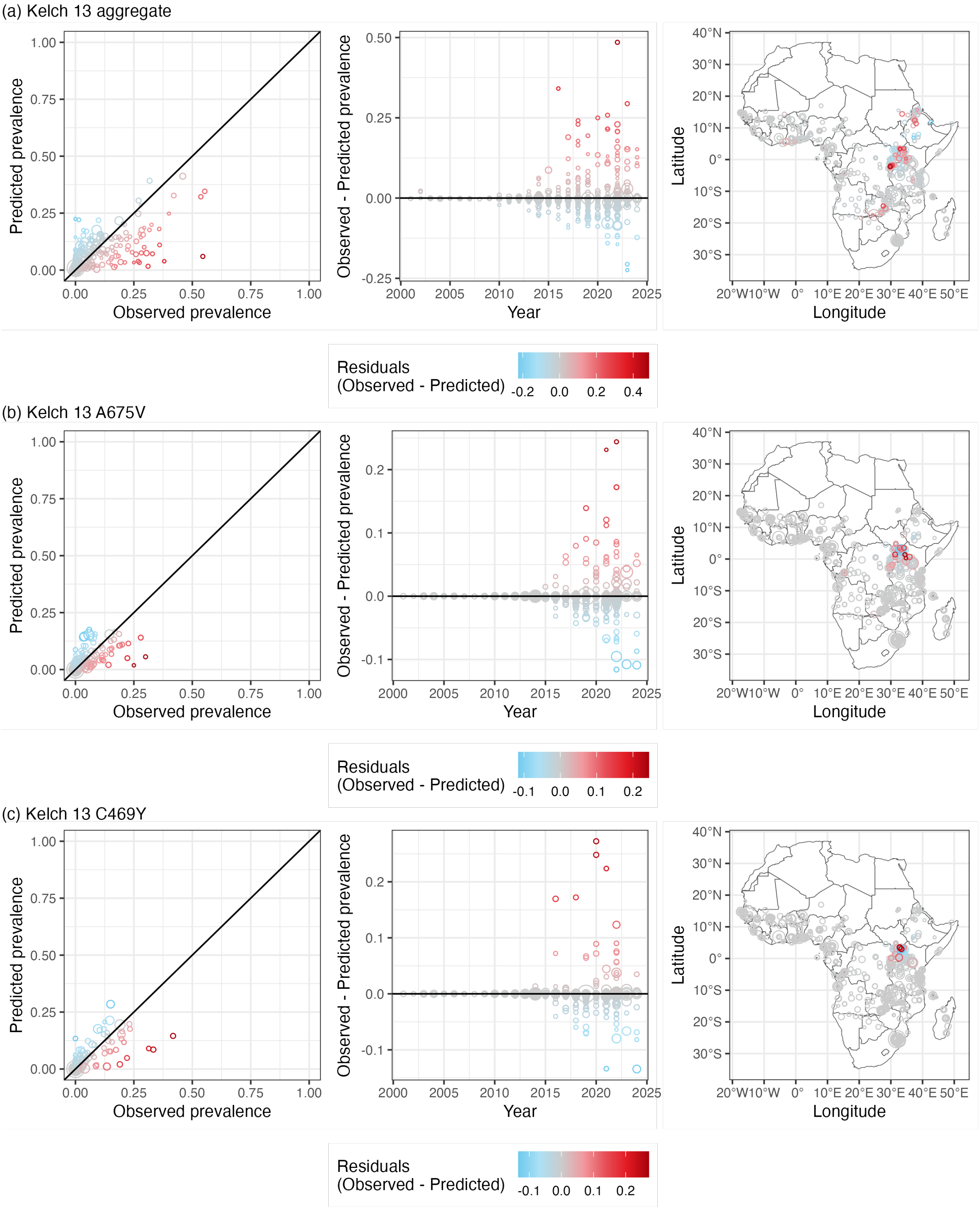


Figure S3: Predicted prevalence over observed prevalence (left), difference in observed and predicted prevalence over time (centre), and difference in observed and predicted prevalence over space (right), for (a) Kelch 13 aggregate model, (b) Kelch 13 A675V model, and (c) Kelch 13 C469Y model.

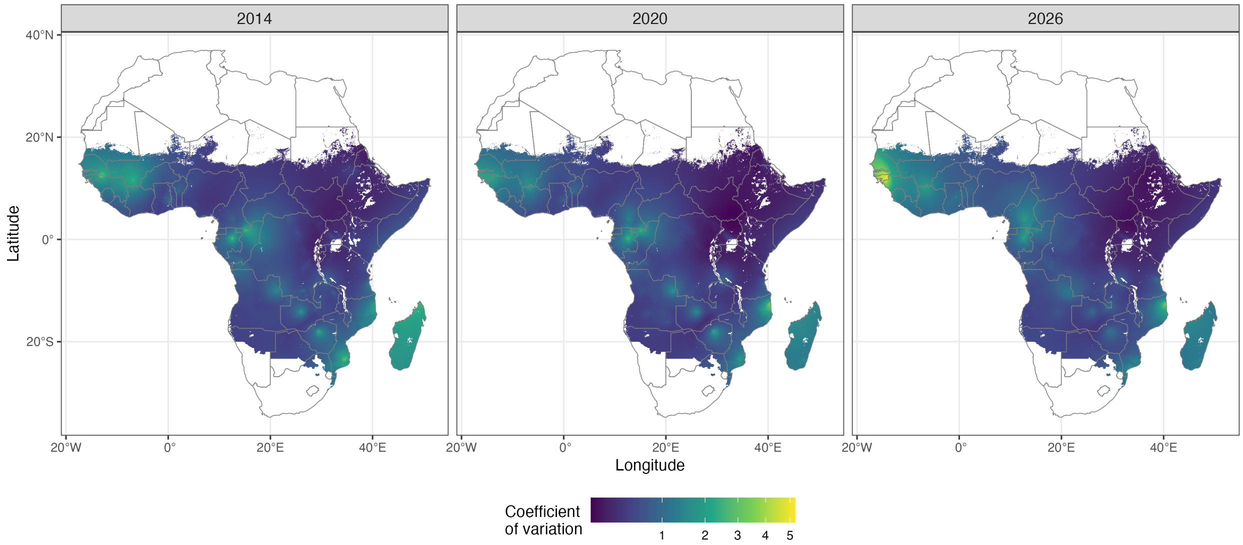


Figure S4: Aggregate Kelch 13 mutation prevalence posterior coefficient of variation: standard deviation of posterior predictions divided by median in 2014 (left), 2020 (centre), and 2026 (right).

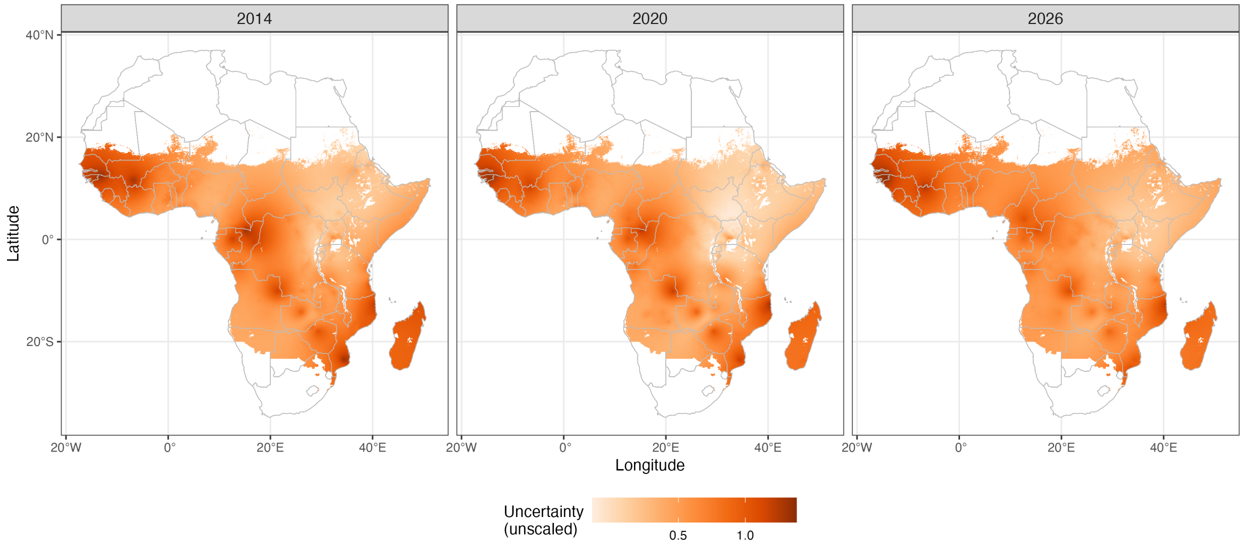


Figure S5: Standard deviation of aggregate Kelch 13 mutation prevalence posterior predictions before samples are transformed to the probability scale in 2014 (left), 2020 (centre), and 2026 (right).

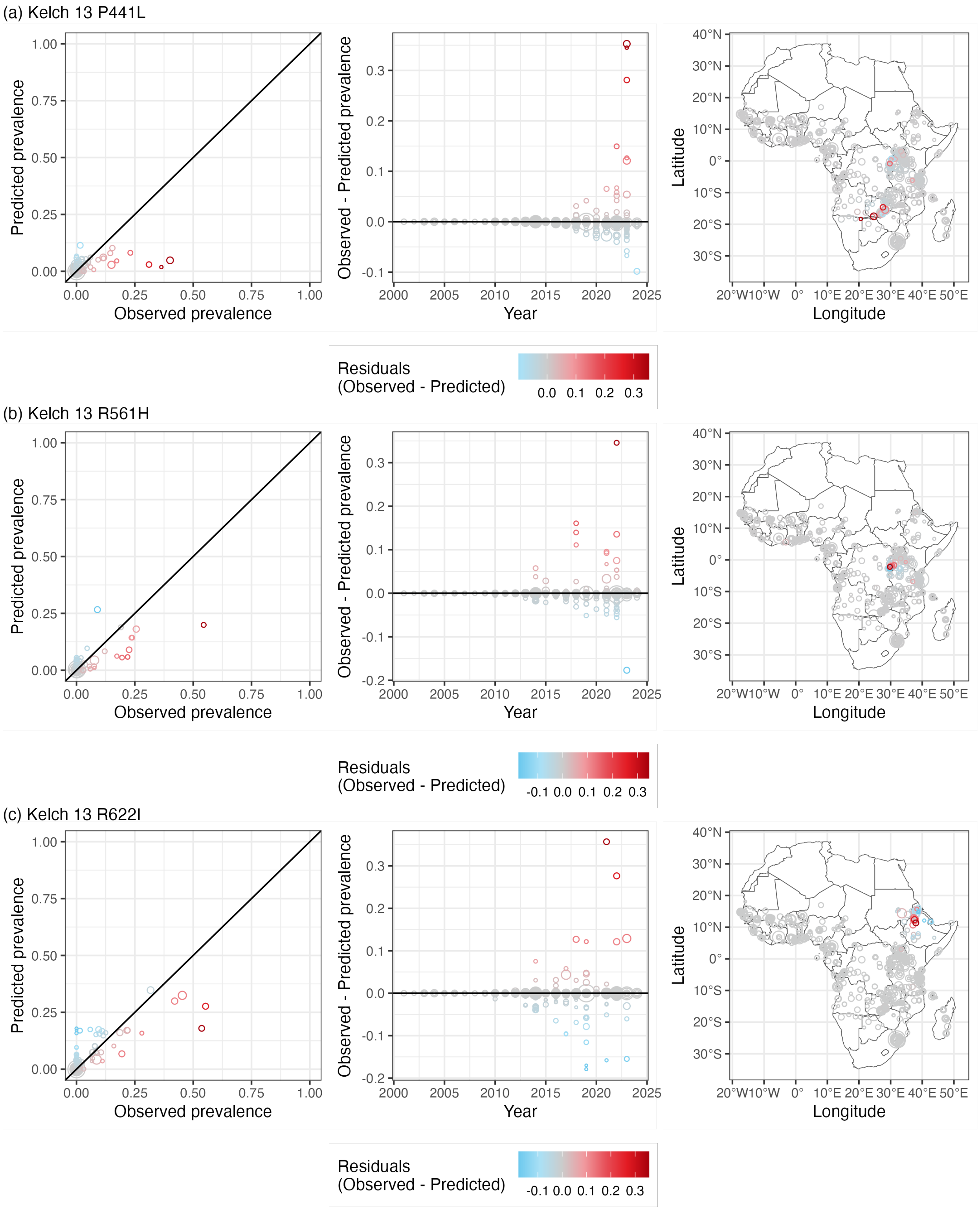


Figure S6: Predicted prevalence over observed prevalence (left), difference in observed and predicted prevalence over time (centre), and difference in observed and predicted prevalence over space (right), for models of (a) Kelch 13 P441L, (b) Kelch 13 R561H, and (c) Kelch 13 R622I.

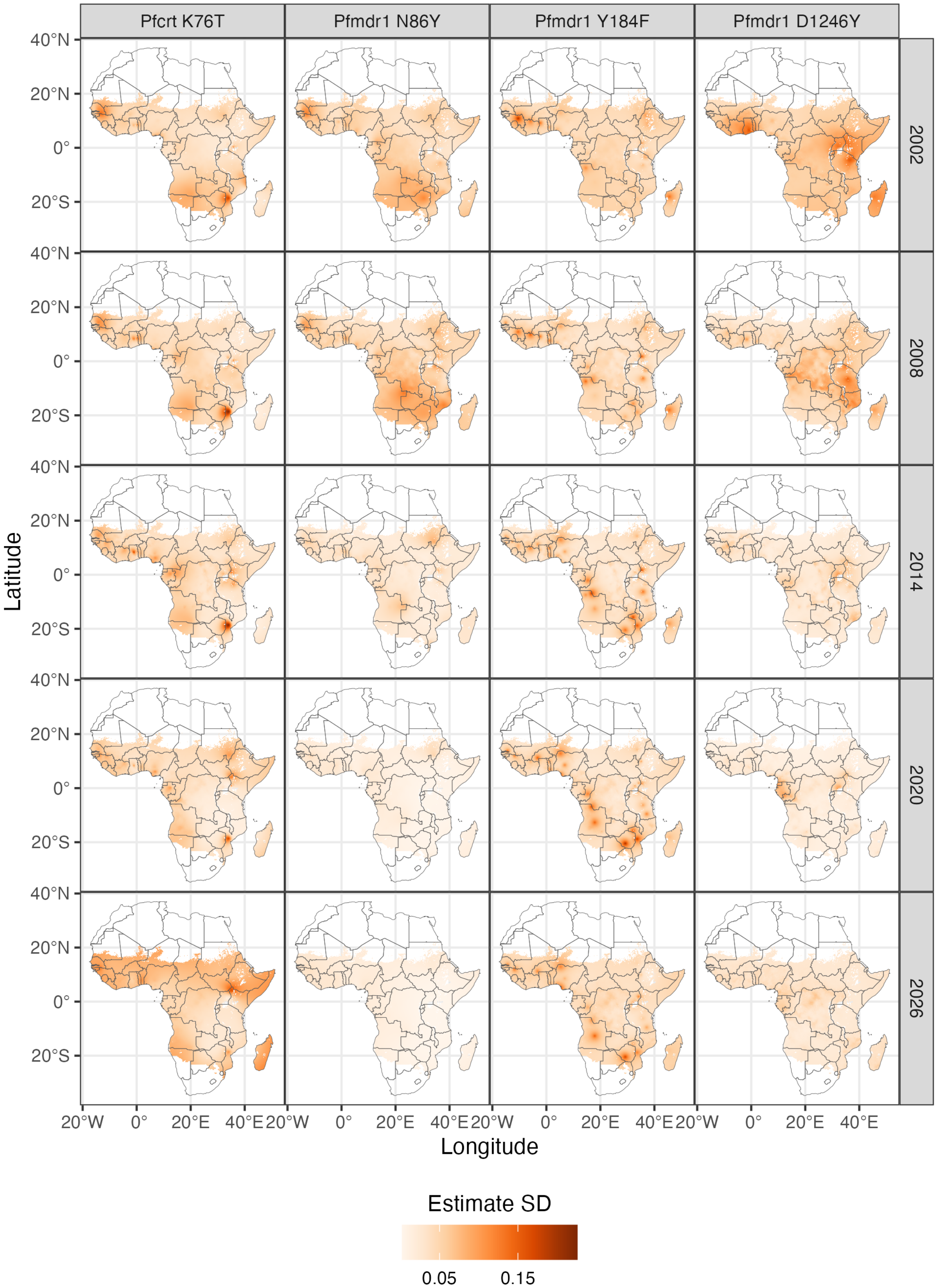


Figure S7: Sample standard deviation of posterior partner drug marker prevalences, corresponding to median estimates in Figure 5. Sample standard deviations before model estimates are logit-transformed to the probability scale are shown in Figure S8.

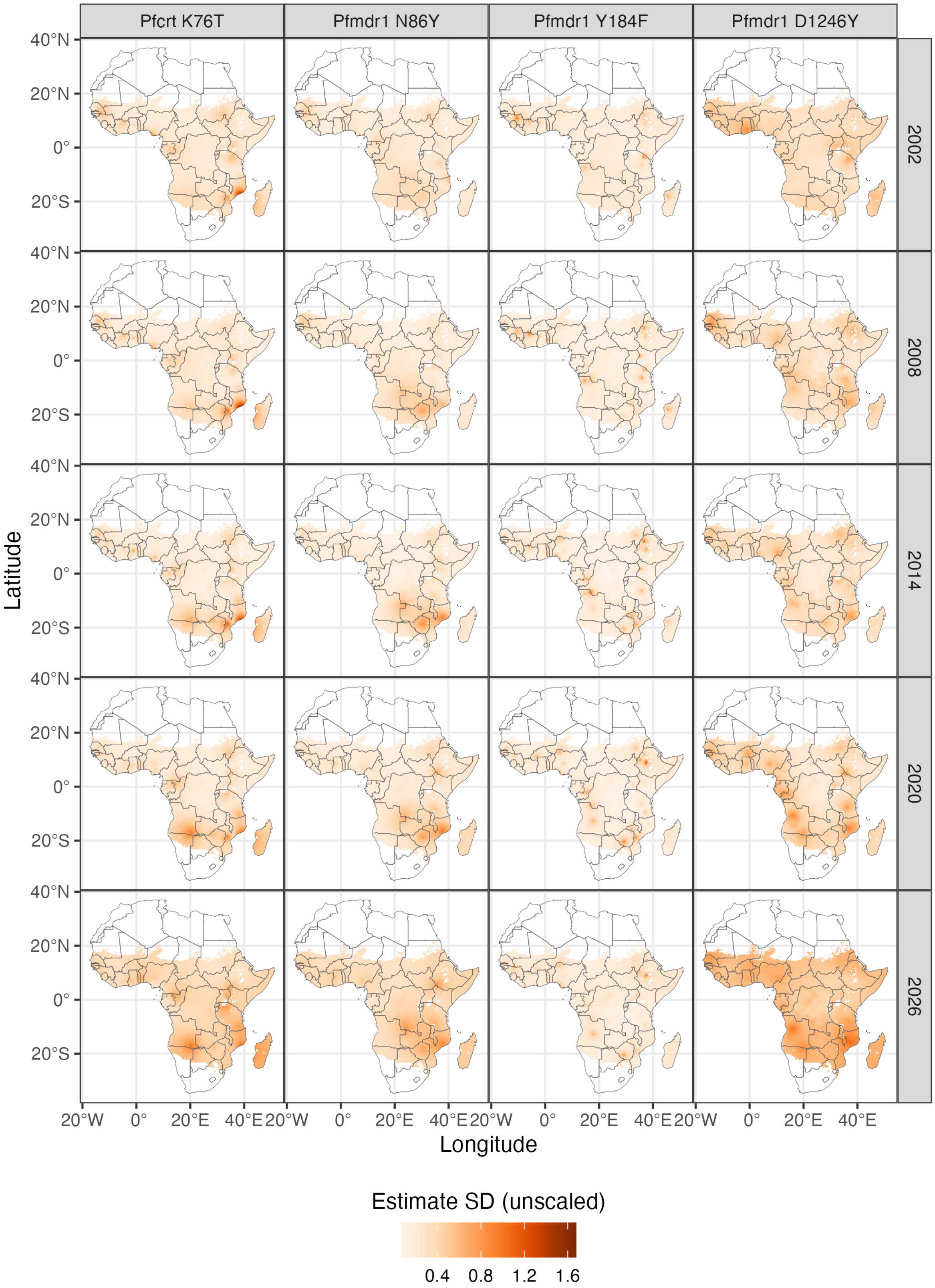


Figure S8: Sample standard deviation of posterior partner drug marker prevalences, before model estimates are logit-transformed to the probability scale (Equation (S3)).

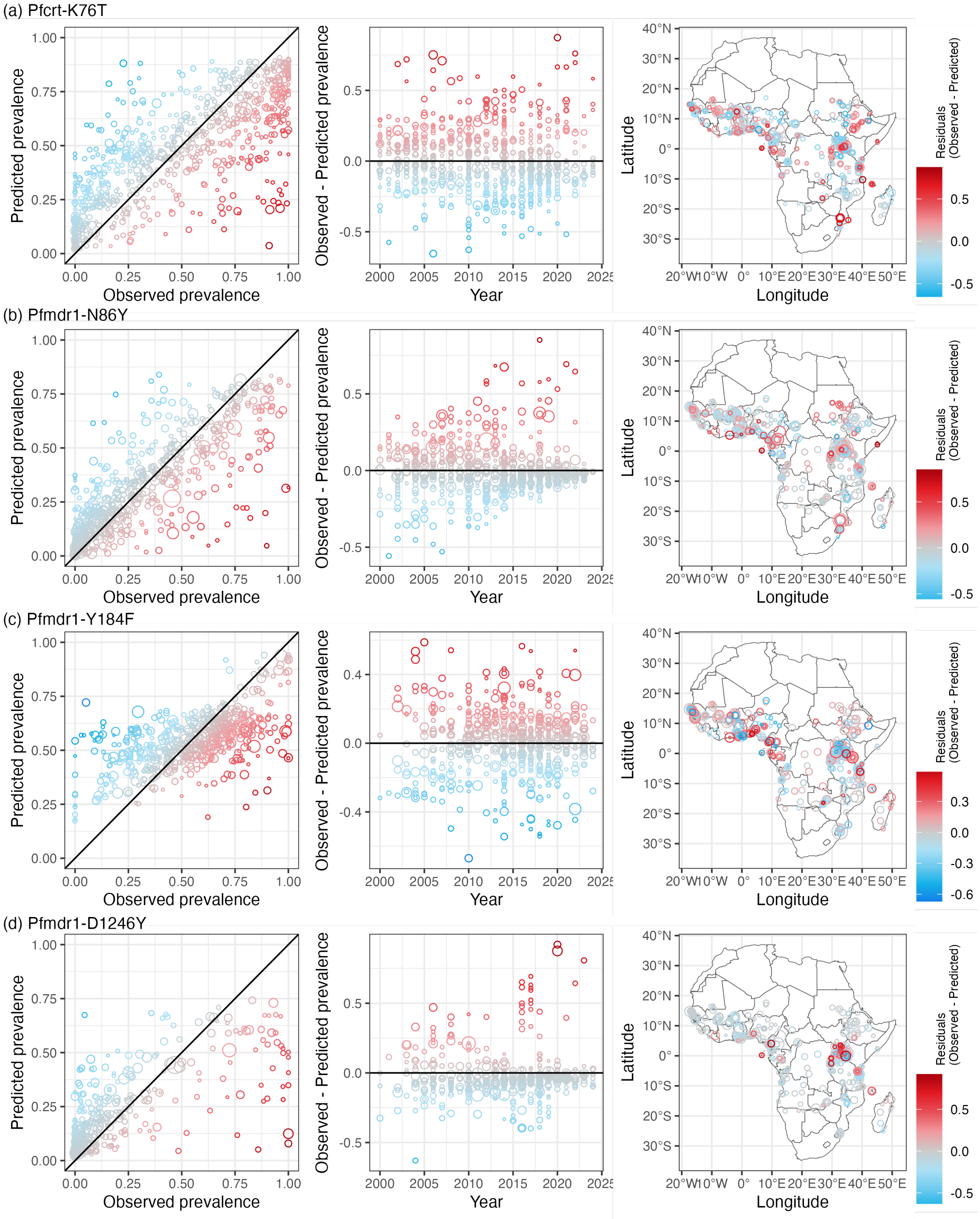


Figure S9: Predicted prevalence over observed prevalence (left); difference in observed and predicted prevalence over time (centre); and difference in observed and predicted prevalence over space (right), for models of estimated prevalence for (a) *Pfcrt*-76, (b) *Pfmdr1*-86, (c) *Pfmdr1*-184, and (d) *Pfmdr1*-1246.

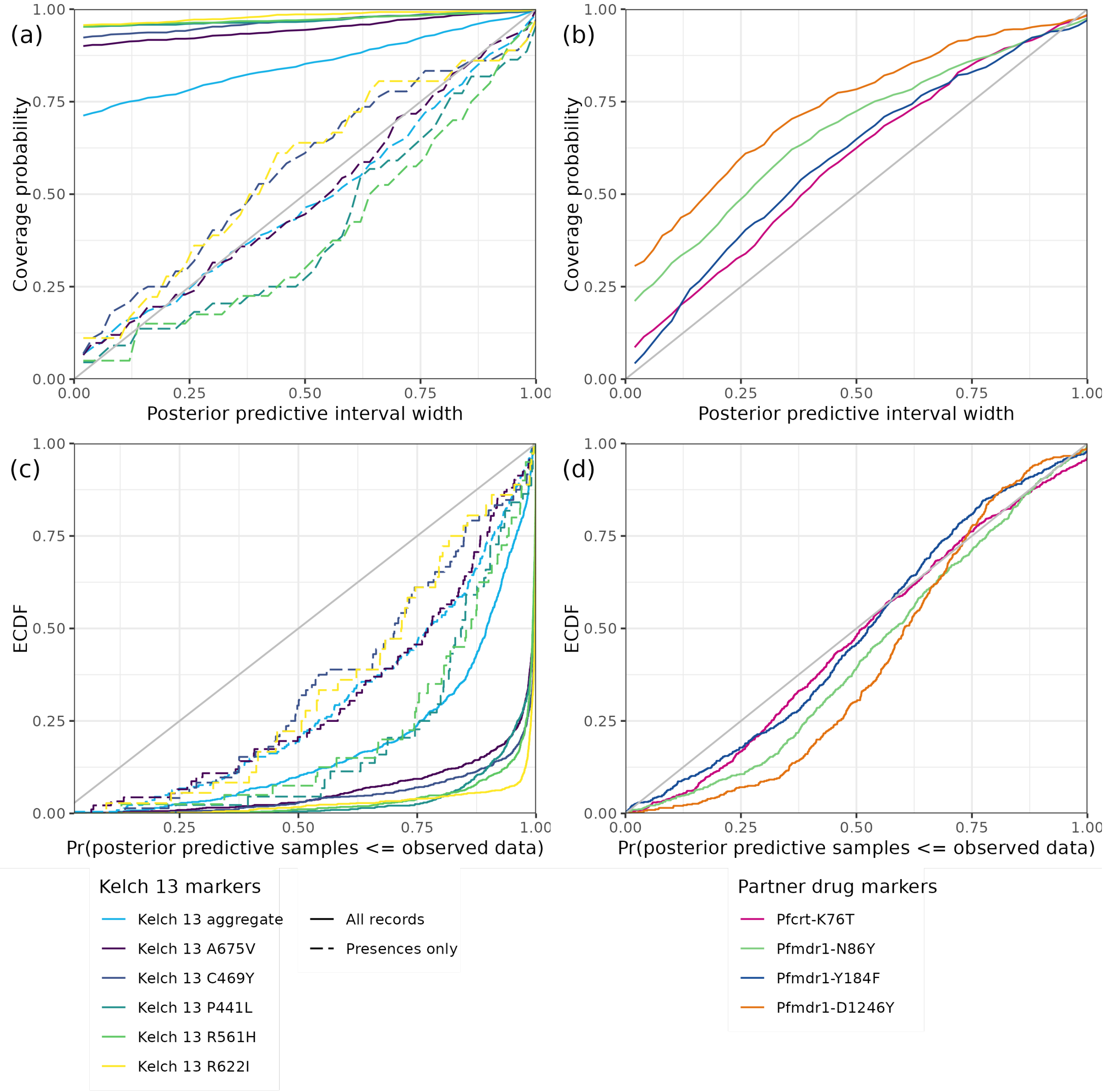


Figure S10: Top: coverage probabilities, or the proportion of prevalence observations covered by posterior credible intervals, for (a) Kelch 13 models and (b) partner drug marker models, given predictions passed through the beta-binomial observation model, with posterior samples for $\rho$, at sites associated with observations in the molecular surveillance dataset. Bottom: probability integral transform empirical cumulative distribution functions (PIT-ECDFs) for (c) Kelch 13 models and (d) partner drug marker models. Dashed lines for Kelch 13 models indicate coverage probabilities and PIT-EDCFs calculated excluding zero-prevalences (or absences of mutants) in the surveillance dataset.

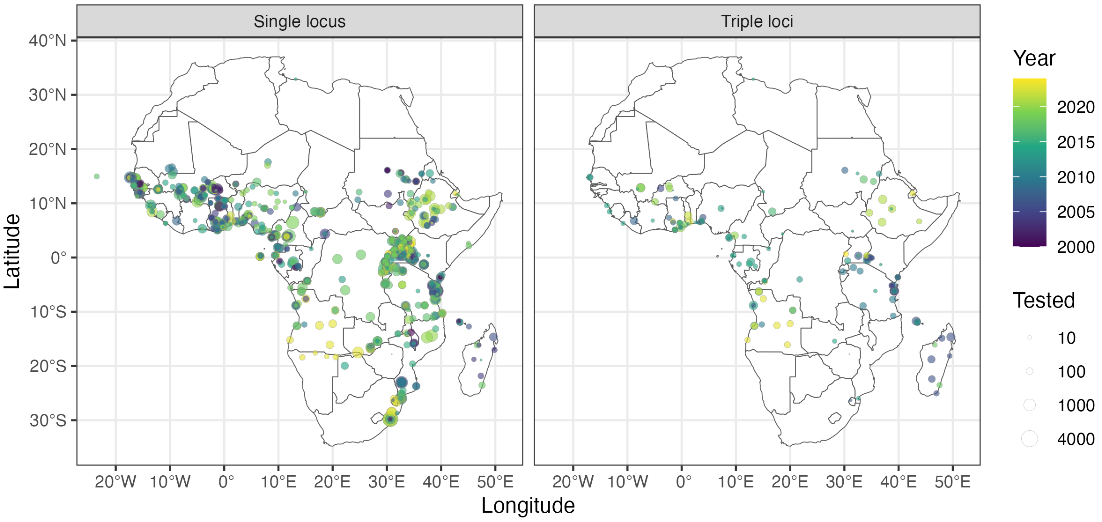


Figure S11: Comparison of distribution of *Pfmdr1* records by publications that reported single locus prevalences (left) and publications that reported haplotype (triple-loci) prevalences (right). Point sizes indicate sample sizes.
