## Supplementary methods for "Estimating the changing prevalence of molecular markers of artemisinin partial resistance in *Plasmodium falciparum* malaria in Sub-Saharan Africa"

In this work we model the prevalence of a shortlist of *P. falciparum* mutations, as proxy for the prevalence of antimalarial resistance in the parasite population, in light of the wider availability and inter-comparability of parasite sequencing data, relative to therapeutic efficacy data [1, 2].

#### S1 Molecular marker prevalence dataset

The data of genomic surveillance of *P. falciparum* we use in this work are largely from the IDDO/WWARN Molecular Surveyor dataset, a living systematic review of published observations of molecular markers for drug resistance in *P. falciparum*. The Surveyor dataset collects published observations of genotyped *P. falciparum* in the population (i.e., in the absence of or before specific treatment or chemoprevention), associated with, at minimum, a country and a three-year sample collection window. The protocol for Surveyor data extraction has previously been published [3]. For Kelch 13, we also include genomic surveillance data from the MARC SE-Africa artemisinin resistance dashboard [4], which collates further published or unpublished surveillance of Kelch 13 in an equivalent format to the IDDO/WWARN Surveyor, with a specific focus on southern and eastern Africa. Where surveillance data was associated with a time window of greater than one year, a single (median) year was imputed.

Data from the World Health Organization (WHO) Malaria Threats Map [5] is included into the MARC SE-Africa dataset. We exclude data from the MalariaGen project on the grounds that the intersection between MalariaGen and the IDDO/WWARN Surveyor is uncertain: the same data may have been published in other articles included in the Surveyor as well as being part of the wider MalariaGen project.

We truncate the dataset to samples collected between 2000 and 2024 (the most recent year in the dataset). Mixed infections are recorded inconsistently in the literature, and thus are recorded in slightly different formats in the Surveyor dataset: because mixed infections are not always separated from mutants (or wildtypes) in the literature, here, for consistency, we group them with the mutants where they have been reported (and subsequently entered into the dataset) separately. We discuss data organisation for each of the models in detail in the following sections.

##### S1.1 Kelch 13

As of February 2026, the IDDO/WWARN Surveyor dataset collates the results of 230 published studies of Kelch 13 surveillance in *P. falciparum* in Africa published between 2014 and 2026. These surveillance data span the years 1995–2024, subsequently truncated to 2000–2024, which excludes one study. The dataset reflects surveillance in 90,881 malaria patients. The MARC SE-Africa dashboard adds 29 peer-reviewed and non-peer-reviewed sources of surveillance data. The addition of the MARC SE-Africa dashboard data increases the full dataset's temporal coverage post-2021 and its spatial coverage of southern and eastern Africa, where the MARC SE-Africa consortium's efforts are focused. The joint dataset reflects surveillance in 118,786 malaria cases (of which 111,454 were Kelch 13-wildtype infections, 2,960 infections had an ART-R-validated mutation, and 420 had an ART-R candidate mutation). (During data extraction for the Surveyor, and subsequently for the MARC SE-Africa dataset, Kelch 13 wildtypes are defined as those where surveillance of the entire Kelch 13 propeller region (loci$>$440) was sequenced and *no non-synonymous mutations* were identified [3].) Of the 20 Kelch 13 mutations defined by the WHO as validated or candidate markers of ART-R in Table S1, 16 appear in the Kelch 13 surveillance dataset and 11 were recorded in 10 or more malaria cases (Figure 1). The five most frequently identified mutations were C469Y ($n = 862$), A675V ($n = 812$), R622I ($n = 723$), R561H ($n = 437$), and P441L ($n = 296$): we examined the spatiotemporal distributions of each of these markers with individual models.

To create input datasets for models of individual Kelch 13 mutations, the dataset was filtered to locations where each of the target mutations were sequenced, with the number of samples with the mutation greater than or equal to zero. For each mutation, a second subset of data was then added: of the set of Kelch 13 wildtypes, those wildtypes recorded at a location in time and space *not* in the set of sites where each of the mutations was already indicated were added to the filtered dataset with the number of cases involving the mutant assigned to zero. From this joint set of presences and absences we therefore have a set of sites associated with a number of malaria patients tested, and a number of patients with each of the Kelch 13 mutations of interest.

To create an input dataset for an aggregate model of all ART-R validated and candidate mutations, the joint Surveyor and MARC SE-Africa dataset was filtered to detections of Kelch 13 mutations associated with ART-R (Table S1). This filtered dataset was aggregated by location, year, and study, to create a site-level number of total mutant isolates with a common sample size (conservatively using the maximum of the set of sample sizes, where there was more than one, to prevent over-estimating surveillance efforts where the same malaria cases are represented in multiple tests for multiple Kelch 13 mutations). This set of mutants was subsequently aggregated to a unique set of locations and years, with a single, summed number of mutant cases and a single sample size. We used as absences the set of locations where wildtypes were identified that were *not* in the set of presences. Putting these two subsets together, we have a set of sites associated with a number of malaria patients tested, and a number of patients with *P. falciparum* including *any* of the mutations included in Table S1.

#### S1.2 Partner drug markers

As of February 2026, the IDDO/WWARN Molecular Surveyor dataset documents 426 publications describing surveillance of *Pfcrt*-K76T in 105,070 malaria patients surveilled between 1978 and 2024. Following aggregation of mixed infections with mutant infections, data are numbers of both mutant and mixed infections associated with sample size, an approximate year, an approximate location[1], and publication. Data were then grouped across multiple publications, to a summed number of mutant/mixed infections with summed sample size, at a set of unique locations in space and time.

The Molecular Surveyor also contains 420 publications describing surveillance of one or more of the markers *Pfmdr1*-86, -184, or -1246 in Africa (of which 103 *do not* describe surveillance of *Pfcrt*-K76T), in 120,059 malaria patients, surveilled between 1978 and 2024. The majority of data of *Pfmdr1*-86, -184, or -1246 surveillance in the Surveyor was extracted at the level of single loci (157,412/162,765 tests; at 3,867/4,587 sites total; Figure S11). Complete haplotype data are often not reported in research articles so are not included in the Surveyor, as data extraction for the Surveyor relies on the information reported in source literature *only*, even though authors reporting single-locus data across all three loci may have had access to complete haplotype data. We note that reporting full haplotypes would be better practice [6] as it would enable meta-analysis and modelling efforts to account for individual-level correlations between each of the mutations involved in the haplotype. Some *Pfmdr1* Surveyor data describes triple haplotypes (including quintuple haplotypes for the additional loci 1034 and 1042); triple-locus data was disaggregated to single-locus data at the site level. At sites where raw single-locus prevalence was not otherwise available for a given locus, the disaggregated triple-locus data was added to the dataset[2]. This cleaning step avoided including the same data twice where both single-locus and triple-locus data was added to the Surveyor for the same study.

Recorded surveillance of the target partner drug markers was most intense in the Lake Victoria region (Figure 4; Figure S1). In the beginning of the study period (post-2000), target partner drug marker prevalences were generally greater than 0.5 (i.e., in favour of the mutant allele) for *Pfcrt*-76 and *Pfmdr1*-86 (Figure 4). By the end of the study period (2026), the prevalence of the mutant genotype of both of these markers was lower throughout Africa, with the exception of the horn of Africa and western Africa, where *Pfcrt*-76T prevalences of 90–100% were sustained. The prevalence of *Pfmdr1*-184F (for which the relationship between mutation status and parasite susceptibility to artemether-lumefantrine (AL) and artesunate-amodiaquine (ASAQ) is inverse to that of the other three modelled partner drug markers) was moderate at the beginning of the study period. By the end of the study period, higher prevalences of the mutant allele *Pfmdr1*-184F, selected by treatment with AL, were observed in the horn of Africa and western Africa (Figure 4). Before 2000, the prevalence of *Pfmdr1*-1246Y was greater than 0.5 in eastern Africa, especially in the Lake Victoria region. By 2026, the prevalence of the mutant genotype (1246Y) was near zero (value) throughout the area of endemic malaria transmission.

[1]Location precision varied from country-level to health district-level.

[2]Sample sizes were not included in site matching. For example, if data of the mutation N84Y and the haplotypes N84-186F-D1246 and N84-Y186-D1246 were available for a site, the record of N84Y was retained and records of Y186F and D1246Y, with associated sample sizes, which may be different from that of N84Y, are appended from frequencies of haplotypes.

### S2 Modelling framework

In this work, we create ten models of markers of antimalarial drug resistance independently: aggregate Kelch 13 mutation prevalence (grouping together the markers associated with ART-R listed in Table S1), the prevalences of the individual Kelch 13 mutations C469Y, A675V, R622I, P441L, and R561H, and the prevalences of five putative markers of reduced susceptibility to artemisinin combination therapy (ACT) partner drugs, *Pfcrt*-K76T, *Pfmdr1*-N86Y, *Pfmdr1*-Y184F, and *Pfmdr1*-D1246Y. We model marker prevalences with Gaussian Process (GP) regression [7]. GPs are able to fit flexible responses with minimal model specification and, if fit within a Bayesian framework, prior beliefs about parameter values can be included in posterior estimates [8]. GPs have been widely used for modelling genetic diversity in *P. falciparum* [9, 10, 11, 12].

The number of malaria patients with a target mutation (or *any* target Kelch 13 mutation, for the Kelch 13 aggregate model) at every location $i$ in time, $t_i$, and space, $x_i$, is often assumed to follow a binomial distribution [9, 12], however in this work we use a beta-binomial observation model to account for possible overdispersion. Where the usual parameterisation of the beta-binomial distribution is Betabinomial$(n, \alpha, \beta)$, we let:

$$p = \frac{\alpha}{\alpha + \beta} \quad \text{and} \quad \rho = \frac{1}{\alpha + \beta + 1}, \tag{S1}$$

where $p$ is the prevalence parameter (i.e., the probability of sampling a malaria patient with the target marker), and $\rho$ is the dispersion parameter. It follows that:

$$\frac{n\alpha}{(\alpha + \beta)} = np \quad \text{and} \quad \frac{n\alpha\beta(\alpha + \beta + n)}{(\alpha + \beta)^2(\alpha + \beta + 1)} = np(1 - p)(1 + (n - 1)\rho) \tag{S2}$$

are the mean and variance of the beta-binomial distribution, respectively. The additional free parameter, $\rho$, models the "intra-cluster" variance, or the extent to which variance in surveillance data is greater than (overdispersed) or less than (underdispersed) the variance of a binomial distribution, $np(1 - p)$.

We assume that prevalence, $p(x_i, t_i)$, of each beta-binomial random variable is itself, once inversely *logit*-transformed, drawn from a latent (or unobserved) normal distribution, $z$. Together, infinitely many $z$ form a GP. GPs consist of a mean function, $\mu(x, t)$ and a covariance function or kernel, $C(x, t)$, parameterised by $\boldsymbol{\theta}_m$ and $\boldsymbol{\theta}_c$, respectively. Hence, we set the number of malaria patients with a target mutation or marker at location $i$ to be:

$$\begin{aligned} N_i^+ &\sim \text{BetaBinom}(n_i, p(x_i, t_i), \rho), \\ \text{logit}(p(x_i, t_i)) &= z(x_i, t_i), \\ z(x, t)|_{\boldsymbol{\theta}_m, \boldsymbol{\theta}_c} &= GP(\mu(x, t), C(x, t)), \end{aligned} \tag{S3}$$

where $n_i$ is the number of malaria cases sampled at location $i$. We allow the mean function of the GP to vary with respect to time (year, $t$, rescaled before model fitting) and estimated *P. falciparum* parasite rate (PfPR; $m(\cdot)$):

$$\mu(x, t) = \beta_0 + \beta_1 t + \beta_2 m(x, t). \tag{S4}$$

We accessed annual rasters of PfPR from the Malaria Atlas Project (MAP) at a resolution of 0.0417° (≈5 km) for the years 2000–2024 [13, 14].

The kernel, $C(x, t)$, of a GP describes the covariance in the target distribution between two points in terms of the distance between the two points. For a spatiotemporal model, this distance is expressed in terms of both space and time. We use a Gneiting class spatiotemporal kernel [10, 15]:

$$C(x_i, t_i, x_j, t_j) = s^2 \left(1 + \frac{(t_i - t_j)^2}{l_t^2} + \frac{\delta_{\text{GC}}(x_i, x_j)}{l_s}\right) + \sigma^2 \mathbb{1}(i = j), \tag{S5}$$

with spatial and temporal lengthscales $l_s$ and $l_t$, nugget variance, $s^2$, and a white noise term with variance parameter $\sigma^2$, to capture within-site variance. Spatial distance is expressed using the great circle distance between two points, $\delta_{\mathrm{GC}}(x_i, x_j)$—this distance function accounts for the curvature of the Earth, which is particularly relevant to continental-scale modelling [10].

We set the priors of the kernel hyperparameters, $l_s, l_t, s$, and $\sigma$ to be weakly informative normals truncated at zero, the priors of mean function coefficients to be weakly informative normals centred at zero, and the prior of the overdispersion parameter, $\rho$, to be log-normal:

$$\begin{aligned} l_s, l_t, s &\sim \mathrm{TruncNorm}(0, 3) \\ \sigma &\sim \mathrm{TruncNorm}(0, 2) \\ \beta_0, \beta_1, \beta_2 &\sim \mathrm{Normal}(0, 3) \\ \rho &\sim \mathrm{LogNorm}(0, 1). \end{aligned} \tag{S6}$$

#### S2.1 Inference

The models are implemented in R using the `greta` package for Bayesian modelling and Markov chain Monte Carlo (MCMC) simulation, and its extension for GPs, `greta.gp` [16]. We adapt `greta.gp` to include spatiotemporal Gneiting class kernels: code is available on GitHub. Model fitting is completed using `greta`'s implementation of Hamiltonian Monte Carlo (HMC), which uses a leapfrog integrator to sample from the model posterior to reduce correlation between successive samples, compared to a Gaussian random walk as in the Metropolis Hastings MCMC algorithm. For each model, we selected 40 inducing points from the surveillance dataset using the $k$-means algorithm, and carry out inference on the resulting sparse GP [17]. We use this reduced rank approximation to manage model fitting time: for a full GP model, fitting time is $O(n^3)$, where $n$ is the number of records in the dataset used for fitting, whereas for a reduced rank GP fitting time is $O(m^2 n)$, where $m$ is the number of inducing points. Model convergence was assessed via the Gelman-Rubin diagnostic [18].

#### S2.2 Predicting marker prevalence

Following inference, we took 500 samples from the resulting posterior to make 500 nominal $5 \times 5$ km$^2$ gridded predictions of mutation prevalence in Africa, for each of the target markers across the years 2000–2028. We summarised these into pixel-wise median and standard deviation surfaces. We make predictions only to areas of endemic malaria transmission, applying a mask based on the MAP's estimate of PfPR for 2024 [13, 14].

#### S2.3 Model validation

To validate models, we assessed model coverage probabilities. Given zero-inflation (particularly in Kelch 13 data), to estimate coverage probabilities for each model we first passed predicted marker prevalences and posterior samples of the dispersion parameter $\rho$ through the beta-binomial observation model. Taking quantiles of the resulting samples as posterior predictive intervals, we assessed the proportion of observed prevalences covered by predictive intervals. We carried out posterior predictive checks via probability integral transform empirical cumulative distribution functions (PIT-ECDFs) [19] for each model: for well-fitted models, PIT-ECDFs reflect a uniform cumulative distribution function.

To assess model fit and predictive performance, we also performed 10-fold cross-validation. Folds were stratified with respect to observed marker prevalence. We reported summaries of root mean square error (RMSE) and $r^2$ across held-out models and under models fit to the complete datasets: while RMSE describes prediction error on the scale of observations (i.e. prevalence), $r^2$ is a scale-agnostic measure of correlation between observed and predicted prevalences. We also evaluate the RMSE and $r^2$ of a baseline model, defined as the median annual observed marker prevalence. This baseline model is assumed as a reasonable "rough guess" a decision-maker might make for marker prevalence, given our surveillance dataset, at a location where no data have been collected.

All code to fit the models described in this manuscript, make predictions, and generate figures is available on GitHub.

### S3 Incidence-adjusted estimates of marker prevalence

To estimate the proportion of annual malaria cases involving mutant *P. falciparum*, we multiply our annual gridded estimates of marker prevalence by the MAP's annual gridded estimates of *P. falciparum* malaria incidence [13], to produce estimates of the number of cases involving the marker per map pixel per year. We estimate the total proportion of cases involving the marker per year by summing the estimated number of cases affected across all map pixels, and dividing by the sum of the corresponding incidence surface. We use point estimates of annual incidence only: we express uncertainty in terms of the 95% credible interval for marker prevalence from the models described in this work. MAP has released estimates of annual *P. falciparum* incidence up to 2024: we use the 2024 estimate for subsequent years.

### References


[1] Christian Nsanzabana, Djibrine Djalle, Philippe J Guérin, Didier Ménard, and Iveth J González. "Tools for surveillance of anti-malarial drug resistance: an assessment of the current landscape". *Malaria journal* 17:1 (2018), p. 75. DOI: 10.1186/s12936-018-2185-9.

[2] Pierre Gashema, James Kagame, Patrick Gad Iradukunda, Emmanuel Edwar Siddig, Sofonias Kifle Tessema, Merawi Aragaw Tegegne, Mazyanga Lucy Mazaba, Mosoka Fallah, Daniel Ngamije, Jean de Dieu Harelimana, et al. "Mapping *Plasmodium falciparum* mutations in Africa: A critical review of emerging drug resistance and implications for malaria control." *International Journal of Infectious Diseases* (2025), p. 108033. DOI: 10.1016/j.ijid.2025.108033.

[3] Sabina Dahlström Otienoburu, Philippe Guerin, Stephanie van Wyk, Caitlin Richmond, Infectious Diseases Data Observatory, Farhad Shokraneh, and Dhol S Ayuen. "Prevalence Of *Plasmodium falciparum* Molecular Markers Associated With Resistance To Antimalarial Drugs: A Systematic Review And Meta-Analysis". *Open Science Framework* (2025). DOI: 10.17605/OSF.IO/8CD3A.

[4] Stephanie van Wyk, Ishen Seocharan, Eulambius M Mlugu, Dhol S Ayuen, Donnie Mategula, Tikhala Makhaza, James Kiarie, Victor Asua, Jimmy Opigo, Aimable Mbituyumuremyi, et al. "The MARC SE-Africa Dashboard: Joining forces to counteract emerging antimalarial resistance in south and east Africa". *medRxiv* (2025), pp. 2025–01. DOI: 10.1101/2025.01.07.25320158.

[5] World Health Organization. *WHO Malaria Threats Map: molecular marker studies of antimalarial drug resistance*. 2025. URL: https://apps.who.int/malaria/maps/threats (visited on 02/24/2026).

[6] Jonathan J Juliano, Cecile PG Meier-Scherling, Neeva Wernsman Young, George A Tollefson, Sean V Connelly, Jonathan B Parr, Melissa D Conrad, Jacob M Sadler, Christopher M Hennelly, Ashenafi Assefa, et al. "Toward Setting Minimum and Optimal Data to Report for Malaria Molecular Surveillance with Targeted Sequencing: The "What" and "Why"". *The American Journal of Tropical Medicine and Hygiene* 113:6 (2025), p. 1169. DOI: 10.4269/ajtmh.25-0147.

[7] Christopher KI Williams and Carl Edward Rasmussen. *Gaussian processes for machine learning*. Vol. 2. 3. MIT press Cambridge, MA, 2006.

[8] Nick Golding and Bethan V Purse. "Fast and flexible Bayesian species distribution modelling using Gaussian processes". *Methods in Ecology and Evolution* 7:5 (2016), pp. 598–608. DOI: 10.1111/2041-210X.12523.

[9] Jennifer A Flegg, Georgina S Humphreys, Brenda Montanez, Taryn Strickland, Zaira J Jacome-Meza, Karen I Barnes, Jaishree Raman, Philippe J Guerin, Carol Hopkins Sibley, and Sabina Dahlström Otienoburu. "Spatiotemporal spread of *Plasmodium falciparum* mutations for resistance to sulfadoxine-pyrimethamine across Africa, 1990–2020". *PLOS Computational Biology* 18:8 (2022), e1010317. DOI: 10.1371/journal.pcbi.1010317.

[10] Yong See Foo and Jennifer A Flegg. "A spatio-temporal model of multi-marker antimalarial resistance". *Journal of the Royal Society Interface* 21:210 (2024), p. 20230570. DOI: 10.1098/rsif.2023.0570.

[11] Apoorv Gupta, Lucinda E Harrison, Minu Nain, Sauman Singh Phulgenda, Rutuja Chhajed, Roopal S Kumar, Aishika Das, Manju Rahi, Philippe J Guerin, Anup R Anvikar, et al. "Model-guided geospatial surveillance system for antimalarial drug resistance". *PLOS Global Public Health* 6:1 (2026), e0004717. DOI: 10.1371/journal.pgph.0004717.

[12] Neeva Wernsman Young, Cécile PG Meier-Scherling, Gina Cuomo-Dannenburg, George A Tollefson, Sean V Connelly, Jacob Marglous, Isabela Gerdes Gyuricza, Kelly Carey-Ewend, Ronald Kyong-Shin, Zachary R Popkin-Hall, et al. "Mapping the prevalence of molecular markers of *Plasmodium falciparum* artemisinin partial resistance in Africa: a spatial-temporal modelling study". *medRxiv* (2025). DOI: 10.64898/2025.12.22.25342873.

[13] Daniel J Weiss, Paulina A Dzianach, Adam Saddler, Jailos Lubinda, Annie Browne, Michael McPhail, Susan F Rumisha, Francesca Sanna, Yalemzewod Gelaw, Juniper B Kiss, et al. "Mapping the global prevalence, incidence, and mortality of *Plasmodium falciparum* and Plasmodium vivax malaria, 2000–22: a spatial and temporal modelling study". *The Lancet* 405:10483 (2025), pp. 979–990. DOI: 10.1016/S0140-6736(25)00038-8.

[14] Simon I Hay, Carlos A Guerra, Peter W Gething, Anand P Patil, Andrew J Tatem, Abdisalan M Noor, Caroline W Kabaria, Bui H Manh, Iqbal R F Elyazar, Simon Brooker, et al. "A world malaria map: *Plasmodium falciparum* endemicity in 2007". *PLoS medicine* 6:3 (2009), e1000048. DOI: 10.1371/journal.pmed.1000048.

[15] Emilio Porcu, Reinhard Furrer, and Douglas Nychka. "30 Years of space–time covariance functions". *Wiley Interdisciplinary Reviews: Computational Statistics* 13:2 (2021), e1512. DOI: 10.1002/wics.1512.

[16] Nick Golding. "greta: simple and scalable statistical modelling in R". *Journal of Open Source Software* 4:40 (2019), p. 1601. DOI: 10.21105/joss.01601.

[17] Edward Snelson and Zoubin Ghahramani. "Sparse Gaussian processes using pseudo-inputs". *Advances in neural information processing systems* 18 (2005).

[18] Dootika Vats and Christina Knudson. "Revisiting the Gelman–Rubin diagnostic". *Statistical Science* 36:4 (2021), pp. 518–529. DOI: 10.1214/20-STS812.

[19] Teemu Säilynoja, Paul-Christian Bürkner, and Aki Vehtari. "Graphical test for discrete uniformity and its applications in goodness-of-fit evaluation and multiple sample comparison". *Statistics and Computing* 32:2 (2022), p. 32. DOI: 10.1007/s11222-022-10090-6.
