## Supplementary tables for "Estimating the changing prevalence of molecular markers of artemisinin partial resistance in *Plasmodium falciparum* malaria in Sub-Saharan Africa"

Table S1: Kelch 13 mutations defined as *validated* or *candidate* markers of ART-R by the World Health Organization (WHO) [1]. The five most frequently identified mutations in Africa are **bolded**: we model these mutations individually as well as in the Kelch aggregate model.

| WHO validated | | WHO candidate |
|---|---|---|
| F446I | I543T | E252Q |
| N458Y | P553L | **P441L** |
| **C469Y** | **R561H** | C469F |
| M476I | P574L | N537I |
| Y493H | C580Y | G538V |
| G533S | **R622I** | V568G |
| R539T | **A675V** | |

Table S2: Summaries of model performance, presented next to summaries of baseline model performance, "predicting" marker prevalence at all sites equal to annual observed median prevalence. Model performance is evaluated after fitting models to the full dataset *and* following 10-fold cross validation, i.e., separating the dataset into ten subsets stratified by marker prevalence, and for each fold training models on 90% of the dataset and making predictions to the withheld 10% of the dataset. RMSE = root mean square error, calculated on the prevalence scale, $r^2$ = coefficient of determination. *Baseline model predictions (annual median observed prevalences) were zero for all years for the P441L and R622I models, which causes the denominator of the correlation coefficient (as calculated by R's `cor()` function) to be zero.

| | Baseline | | Spatiotemporal GP | | | | | |
|---|---|---|---|---|---|---|---|---|
| | | | Full dataset | | 10-fold holdout | | | |
| | | | | | RMSE | | $r^2$ | |
| | RMSE | $r^2$ | RMSE | $r^2$ | Mean | SD | Mean | SD |
| Kelch 13 aggregate | 0.074 | 0.054 | 0.048 | 0.562 | 0.050 | 0.010 | 0.528 | 0.085 |
| Kelch 13 A675V | 0.031 | 0.015 | 0.020 | 0.545 | 0.020 | 0.006 | 0.564 | 0.232 |
| Kelch 13 C469Y | 0.034 | 0.042 | 0.021 | 0.612 | 0.020 | 0.007 | 0.724 | 0.181 |
| Kelch 13 P441L | 0.025 | * | 0.021 | 0.281 | 0.019 | 0.013 | 0.279 | 0.182 |
| Kelch 13 R561H | 0.027 | 0.000 | 0.017 | 0.613 | 0.030 | 0.044 | 0.502 | 0.260 |
| Kelch 13 R622I | 0.040 | * | 0.023 | 0.673 | 0.020 | 0.011 | 0.740 | 0.216 |
| *Pfcrt*-K76T | 0.295 | 0.299 | 0.218 | 0.598 | 0.197 | 0.021 | 0.673 | 0.073 |
| *Pfmdr1*-N86Y | 0.192 | 0.556 | 0.152 | 0.707 | 0.152 | 0.020 | 0.686 | 0.089 |
| *Pfmdr1*-Y184F | 0.222 | 0.069 | 0.184 | 0.349 | 0.184 | 0.011 | 0.355 | 0.113 |
| *Pfmdr1*-D1246Y | 0.249 | 0.143 | 0.170 | 0.565 | 0.171 | 0.038 | 0.571 | 0.122 |
